# Health Response to Problematic Usage of the Internet: A Global Survey on Trends, Available Treatments and Key Challenges

**DOI:** 10.1101/2025.05.20.25327972

**Authors:** Arash Khojasteh Zonoozi, Joe Schofield, Fateme Sadat Abolghasemi, Sophia Achab, Henrietta Bowden-Jones, Zsolt Demetrovics, Mohsen Ebrahimi, Naomi Fineberg, Yasser Khazaal, Hae Kook Lee, Kristiana Siste, Dan J Stein, Anise M.S. Wu, Mehran Zare-Bidoky, ISAM-GEN Societies’ Experts, Marc N. Potenza, Alexander Mario Baldacchino, Hamed Ekhtiari

## Abstract

**Background and Aims:** Problematic usage of the internet (PUI) is a growing concern as technology evolves, with over 5.3 billion individuals, including children, using the internet globally. While specific forms of PUI, such as those involving online gaming and gambling, have been recognized as disorders in major diagnostic manuals, there remains a lack of global data regarding prevalence, treatment and health responses to PUI. This study aimed to examine the magnitude, treatment and health responses to PUI at an international level and identify gaps in knowledge.

**Methods:** We conducted a global survey within the International Society of Addiction Medicine’s Global Expert Network (ISAM-GEN), involving addiction societies from 38 countries across Europe (13), Asia/Oceania (12), the Americas (8) and Africa (5). Response to PUI was assessed across various domains, including non-specific PUI and problematic online gaming, problematic online gambling, problematic online pornography, problematic social media use and problematic online buying/shopping. The survey structure included sections on six case scenarios representing different PUI subtypes, each followed by targeted questions, along with an evaluation of the significance of PUI, country-level health responses to PUI and the perceived severity of specific PUI subtypes.

**Results:** Problematic online gambling (94.8%) and online gaming (86.9%) emerged as the most frequently reported forms of specific PUI, categorized as either frequent or occasional. These were followed by problematic use of social media (84.2%), online pornography (68.3%) and online buying/shopping (52.6%). Psychotherapeutic approaches, such as cognitive behavioral therapy, were identified as the most widely available treatments for PUI, accessible in over 70% of countries surveyed. Despite increasing global attention to PUI, reflected in the establishment of PUI-focused interest groups in 44.7% of the surveyed societies, significant gaps remain. These gaps include the absence of professional certifications, reported by 78.9% of societies, insufficient educational plans for practitioners (68.4%) and a perceived lack of expert training programs (63.2%). Such deficiencies are concerning given that 65.8% of societies emphasized the projected 10-year severity of PUI as either extremely or very important.

**Conclusion:** While not as rigorous as representative community surveys, this survey and its findings highlight the global importance of PUI and suggests critical gaps in healthcare responses. The disparity between awareness of PUI’s significance and the limited resources to address it warrant urgent interventions internationally. Future efforts should focus on enhancing training programs and investing in sustainable solutions to monitor and mitigate the growing burden of PUI worldwide.

## INTRODUCTION

Digital technologies have profoundly transformed modern life, deeply integrating communication, education, work and leisure. While these innovations have facilitated numerous societal advancements, concerns have been raised about their potential adverse effects on mental health (1, 2). Among these concerns, problematic usage of the internet (PUI) has garnered significant attention from researchers, clinicians and policymakers (3, 4). Since the mid-1990s, when internet adoption became widespread in developed countries, its growth has been both rapid and extensive. Today, over 5.3 billion people, representing approximately two-thirds of the global population—including children—use the internet. Social media platforms account for a substantial portion of this digital engagement, with nearly 5 billion individuals actively participating (5).

The prevalence of PUI has become increasingly evident. A 2020 systematic review and meta-analysis estimated that around 7% of the global population shows signs of internet addiction, with rising prevalence estimates over time, especially among younger demographics (6). These findings underscore the growing importance of addressing PUI as a public health issue (7). The Lancet Psychiatry Commission on PUI has emphasized the global significance of PUI, advocating for increased attention and coordinated efforts to address its challenges (8).

PUI may manifest in various forms, some of which are officially recognized in diagnostic manuals. For example, gambling and gaming disorders, each with online specifiers, are categorized primarily as disorders due to addictive behaviors within the International Classification of Diseases 11th Revision (ICD-11) (9). Compulsive sexual behavior disorder, frequently expressed online, is classified as an impulse control disorder (10, 11). Other manifestations include online compulsive buying/shopping, cyberchondria, cyberbullying, problematic use of social media and digital hoarding (3). These varied presentations highlight the complexity of PUI and the need for diverse intervention strategies.

A critical challenge in addressing PUI is the scarcity of comprehensive data on available treatments worldwide. For instance, while some countries have integrated PUI interventions into existing mental health services, others lack dedicated resources entirely (12). Surveys have shown that fewer than 30% of nations have formal guidelines or policies addressing PUI, leaving significant gaps in treatment accessibility and quality (8). This situation hinders the development of informed strategies to address the issue on a global scale and underscores the need for research aimed at mapping treatment landscapes and identifying disparities in access (13).

In alignment with this growing concern about PUI, we conducted a global survey to examine trends, health responses, prevalence and the perceived severity of this phenomenon. This study also surveyed addiction professionals regarding the preparedness of health systems to manage PUI while identifying critical gaps in current practices. The survey utilized the International Society of Addiction Medicine Global Expert Network (ISAM-GEN), a platform connecting addiction medicine societies worldwide. Through this network, diverse perspectives were gathered, offering a comprehensive view of the global PUI landscape (14).

## METHODS

### Participants

The study initially invited participants from the ISAM-GEN database of 48 national addiction medicine societies. Key contacts from the societies (participants) were experienced addiction medicine professionals, serving as representatives of their national addiction medicine societies. Eligibility criteria mandated that participants be active members of their respective societies and actively engaged in clinical, research, or policy-related activities pertinent to addiction medicine. To ensure credibility, participants were either high-ranking members of their societies (e.g., presidents, vice presidents, or section directors/chairs) or individuals nominated directly by such officials to complete the survey on behalf of the society. Recruitment was conducted via direct email invitations, followed by up to seven weekly or biweekly reminders to non-responders to maximize participation.

### Measures

A structured survey instrument was developed and validated by the steering committee, a diverse panel of international experts in PUI who ensured its content validity (see supplementary materials for a list of members). The questionnaire was designed to comprehensively assess various aspects of PUI and consisted of four main sections, as described below.

#### PUI case scenarios

This section included case scenarios drafted based on criteria from major diagnostic manuals such as the DSM-5 and ICD-11 wherever applicable. These scenarios were designed to represent six specific PUI subtypes: non-specific PUI, problematic online gaming, problematic online gambling, problematic use of online pornography, problematic use of social media and problematic online buying/shopping. The scenarios were placed at the beginning of the survey to ensure that respondents had a consistent understanding of each PUI subtype and its definition while answering subsequent sections of the survey. Questions in this section evaluated several aspects, including the prevalence (frequency of each subtype in respondents’ regions), the most likely initial and ultimate providers of treatment, the expected treatment approaches, the anticipated primary treatment settings and respondents’ evaluations of the expertise of the addiction workforce and the health system’s capability to address these scenarios. Full survey questions are available in the supplementary materials.

#### Societal and organizational importance of PUI

Items exploring topics such as the existence of interest groups, expert training programs, certifications, perceived current and future severity of PUI, research development, investment plans and awareness efforts.

#### Country-level health responses

Questions addressing health responses to PUI, including the availability of treatment programs, expert awareness programs, general educational programs, prevention programs, screening programs, community awareness initiatives and the availability of various treatment modalities.

#### Severity of distinct PUI subtypes

Items assessing the perceived severity of specific PUI subtypes. The full survey draft is available in the supplementary materials.

### Procedure

The survey was disseminated online using Qualtrics software, Version August 2024, with distribution occurring over two-and-a-half months (late July 2024 - early October 2024). Invitations were sent via email, accompanied by follow-up reminders to maximize participation. To ensure accessibility, the survey was designed to be mobile-friendly and available in English, the primary language of ISAM-GEN communications. Participation was voluntary and no financial incentives were offered. The survey underwent pre-testing with a small group of ISAM-GEN assistant officers to ensure clarity, relevance and cultural adaptability. All steering committee members approved the questionnaire before its publication.

### Statistical analysis

Data were analyzed using RStudio (version 2024.9.0.375). Descriptive statistics, including frequencies, percentages, mean and standard deviations were calculated to summarize survey responses across all sections including the prevalence of PUI subtypes, organizational and societal importance, country-level health responses and perceived severity of PUI subtypes. The survey form required complete responses for submission. Partial data were only possible if a respondent abandoned the survey before completion, in which case their responses were excluded from analyses.

### Ethics

Ethical approval for the study was obtained from the University of St Andrews School of Medicine Ethics Committee, with approval number MD18020. All participants provided informed consent electronically before beginning the survey. Data confidentiality was protected by anonymizing responses and securely storing data on password-protected servers. Participants were informed of their right to withdraw from the study at any time without consequence.

## RESULTS

### Demographic characteristics

Out of the 48 national addiction medicine societies listed in the ISAM-GEN database, 38 societies responded across Europe (13), Asia (11), South America (5), Africa (5), North America (3) and Oceania (1), resulting in a response rate of 79.2%. Figure 1 depicts the worldwide distribution of countries participating in this survey study. Among the respondents, 81.6% (n = 31) held senior leadership roles, such as president, vice president, or chair/director/board member, within their respective societies. The average length of experience working with individuals affected by behavioral addictions, either directly or indirectly, was 20.7 years (SD = 8.6). More than half of the respondents were affiliated with universities or teaching hospitals (52.6%, n = 20) and most participants were primarily engaged in clinical activities, with 63.2% (n = 24) working as clinicians.

**FIGURE 1.**
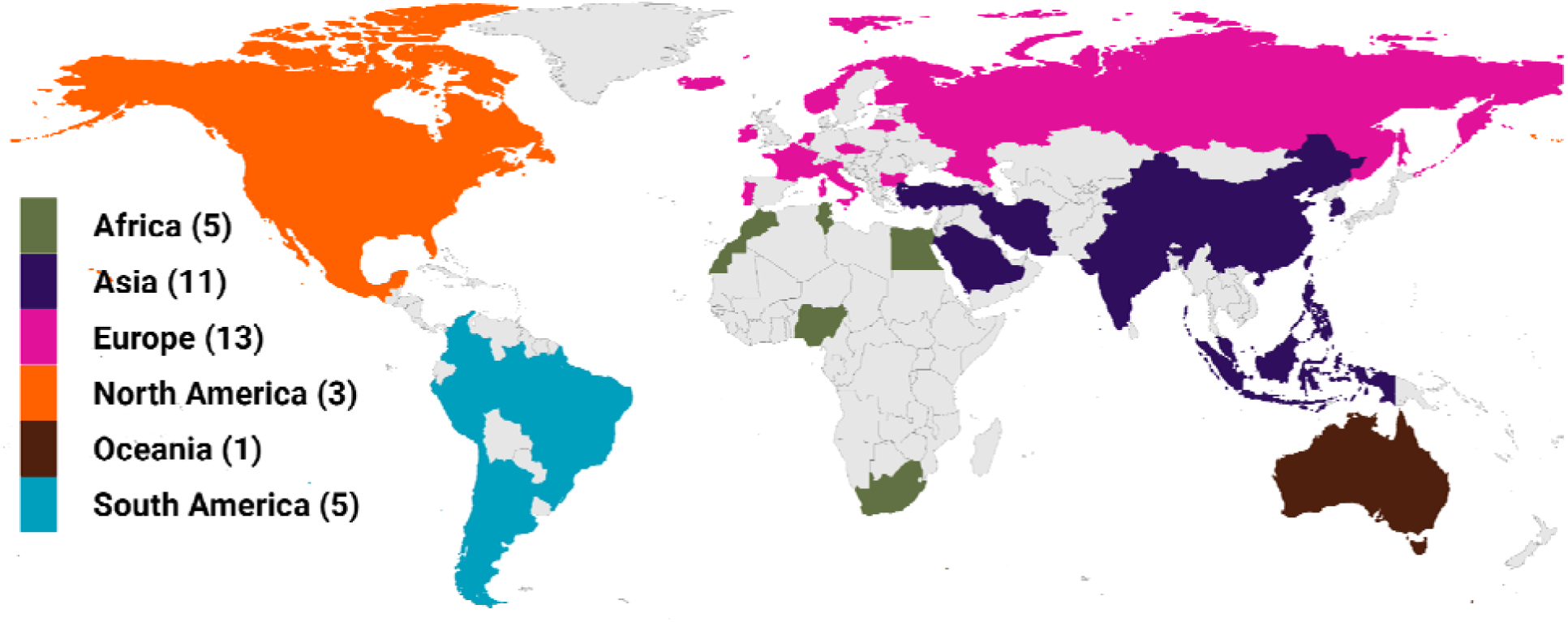
Global distribution of the societies participating in the survey. List of the participating countries is as follows: Argentina, Australia, Brazil, Bulgaria, Canada, Chile, China, Colombia, Czechia, Egypt, Finland, France, Iceland, India, Indonesia, Iran, Ireland, Italy, Lithuania, Malaysia, Malta, Mexico, Morocco, Nepal, Netherlands, Nigeria, Norway, Peru, Philippines, Portugal, Russia, Saudi Arabia, Singapore, South Africa, South Korea, Tunisia, Turkiye and the USA. The numbers in parentheses following the continent names indicate the number of countries from each continent included in this study.

### Prevalence and severity of PUI subtypes

The prevalence of PUI, determined by respondents rating the frequency of each presented scenario on a Likert scale, demonstrated variations across the different subtypes. Non-specific PUI was reported by 89.5% (n = 34) of respondents, summing frequent and occasional cases. Problematic online gambling exhibited the highest prevalence, with 94.7% (n = 36) combining these categories. Similarly, problematic online gaming was reported by 86.8% (n = 33) as frequent or occasional. Problematic online buying/shopping was perceived by 52.6% (n = 20) across these categories, whereas problematic use of social media was noted by 84.2% (n = 32). Problematic use of online pornography was described by 68.4% (n = 26) as frequent or occasional. Figure 2 reflects the general and country-specific prevalence of PUI subtypes.

**FIGURE 2.**
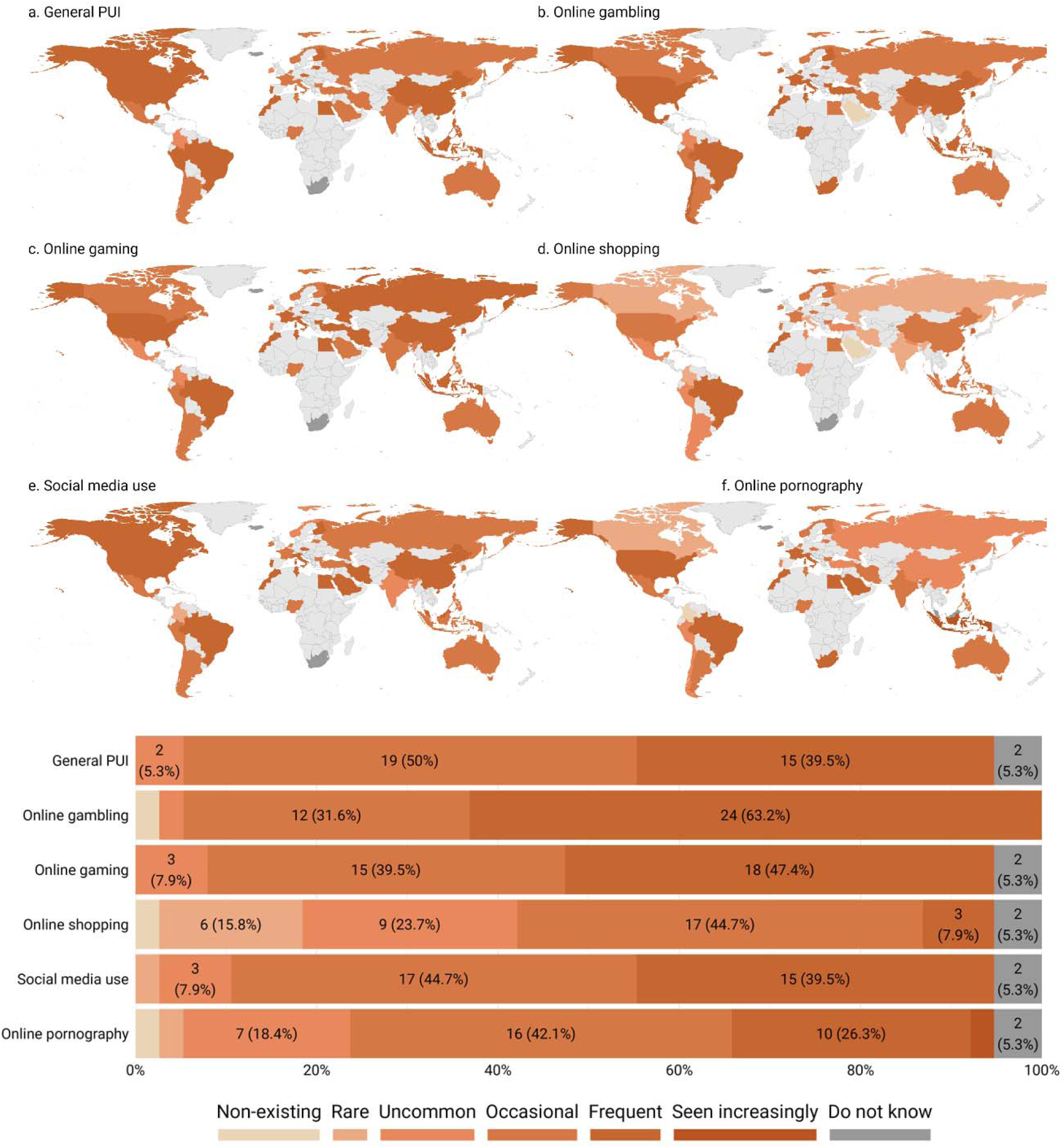
Frequency of distinct PUI branches. The six world heat maps provide a country-specific visualization of PUI branches, using the Likert scale to represent responses ranging from non-existing to seen increasingly. Additionally, the Likert scale diagram illustrates the frequency of these distinct branches as reported by participating societies.

Regarding severity, problematic online gambling stood out, with 10.5% (n = 4) rating it as very severe and 13.2% (n = 5) as severe. Problematic use of social media also had notable ratings, with 18.4% (n = 7) classifying it as severe. Problematic online gaming and use of pornography were rated as severe by 13.2% (n = 5). In contrast, problematic online buying/shopping was rarely classified as severe (2.6%, n = 1), with most respondents rating it as mild (26.3%, n = 10) or very mild (23.7%, n = 9). Moderate ratings were common across subtypes, particularly for problematic online gambling (29.0%, n = 11), online pornography (21.1%, n = 8) and social media (31.6%, n = 12). A substantial proportion of respondents expressed uncertainty about the severity of specific subtypes, particularly for online pornography. Supplementary Figure 1 shows the general and country-specific severity of PUI subtypes.

### Treatment services for PUI

The treatment options available for PUI displayed an almost consistent pattern between PUI subtypes. CBT-based psychotherapy was the most frequently reportedly available treatment, reported for most PUI types, including problematic use of online pornography (81.6%, n = 31), use of social media (79.0%, n = 30), online gambling (76.4%, n = 29), online gaming (73.7%, n = 28), online buying/shopping (71.1%, n = 27) and non-specific PUI (68.4%, n = 26). Psychoeducation was also commonly reported, particularly available for problematic use of online pornography (57.9%, n = 22) and online buying/shopping (52.6%, n = 20). Pharmacological treatments were more frequently reported available for problematic online gambling (52.6%, n = 20) and use of online pornography (29.0%, n = 11). In comparison, mindfulness-based interventions and peer group support therapy were reported more moderately for problematic online gaming (34.2%, n = 13) and use of online pornography (31.6%, n = 12). System-level interventions were less frequently reported available (ranging between 10.5% to 26.3%) and alternative medicine options were rarely considered available (ranging between 2.6% to 5.3%). Figure 3 summarizes the general and country-specific availability of various treatment options for PUI subtypes.

**FIGURE 3.**
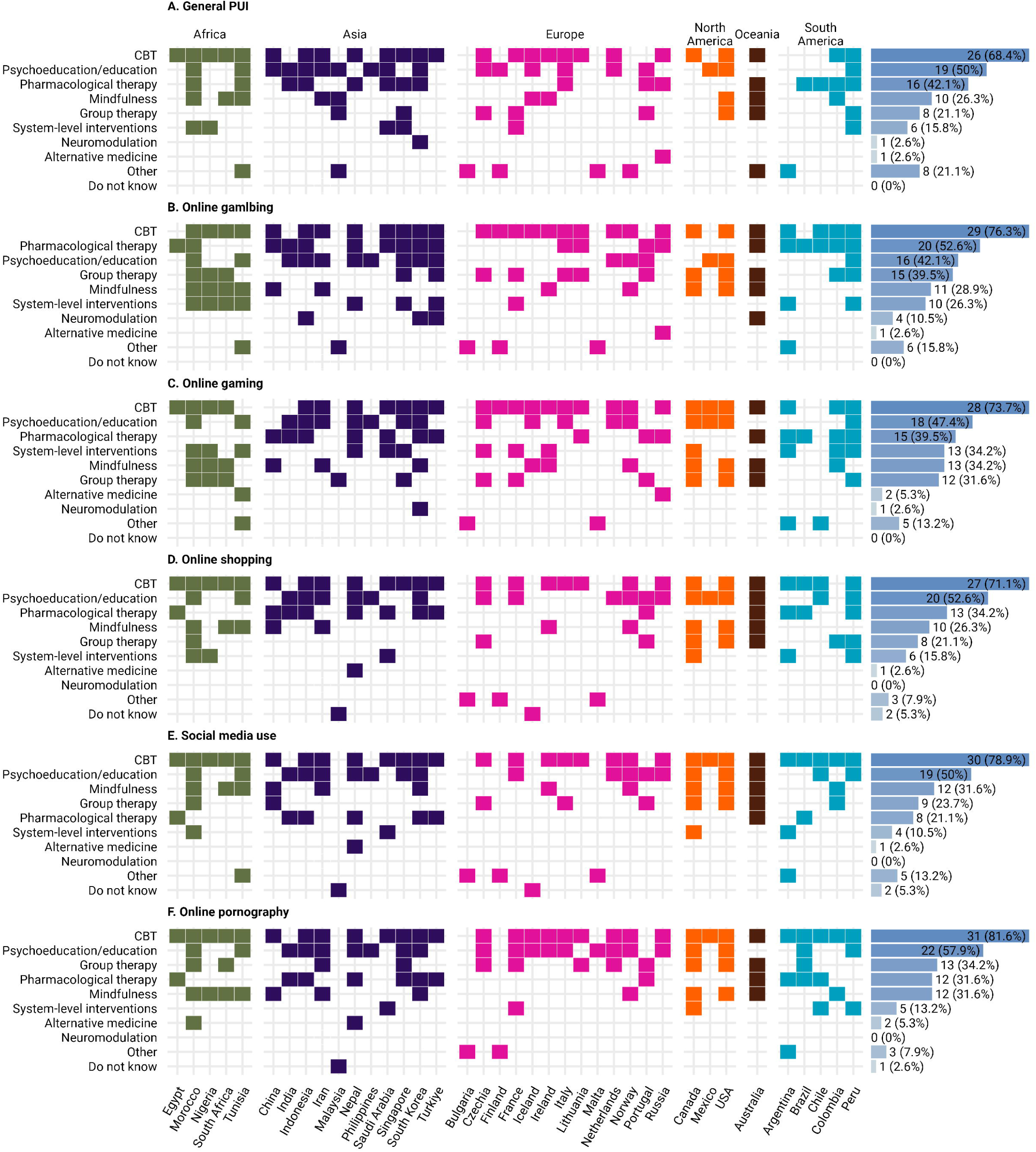
PUI available treatment options for distinct PUI branches. This figure illustrate the availability of various treatment facilities for PUI across different countries. The complete list of treatment options included in the survey, as reported by the respondents, was as follows: Pharmacological therapy, Neuromodulation (different brain stimulations including transcranial magnetic stimulation), Psychoeducation or education only, CBT-based psychotherapy, Mindfulness-based interventions, Peer group support therapy, Alternative medicine interventions (including homeopathy and acupuncture) and System-level interventions (family interventions, school, community, etc.).

Long-term (more than one month) outpatient programs were the most frequently reported available first-line treatment setting for all PUI subtypes, with higher availability for problematic online gambling (63.2%, n = 24), use of online pornography (60.5%, n = 23) and online gaming (55.3%, n = 21). Short-term (up to one month) outpatient programs were also commonly reported to be available as first-line settings, particularly for problematic use of social media (39.5%, n = 15) and online buying/shopping (34.2%, n = 13). Inpatient programs, online long-term programs and other alternative settings were reported rarely, with availability ranging from 0% to 5.3%.

The care providers most frequently reported as the initial point of contact for individuals with different subtypes of PUI were psychologists (42.1% to 50%), followed by general practitioners or family doctors (18.4% to 39.5%). Beyond initial care, follow-on treatment was most commonly provided by psychologists (36.8% to 47.4%) and psychiatrists (29.0% to 36.8%). Addiction medicine specialists were less frequently noted as ultimate care providers (5.3% to 29.0%), while involvement from other health practitioners or emergency doctors was rarely reported (0% to 5.3%). General practitioners were noted as points of initial contact but seldom for ultimate treatment, highlighting the multidisciplinary nature of managing PUI.

### Health response towards PUI

For all PUI subtypes, the evaluation of the addiction workforce’s expertise indicated a predominance of intermediate levels, with most responses categorized as “competent” (29.0% to 39.5%) or “advanced beginner” (26.3% to 36.8%). Higher levels of expertise, such as “proficient” (0% to 15.8%) or “expert” (0% to 10.5%), were reported far less frequently, while novice levels ranged from 13.2% to 26.3%. These results reflect a workforce with foundational skills but limited advanced specialization. In contrast, the capability of the broader health systems to manage PUI scenarios was rated more positively overall. The majority rated the systems as “somewhat capable” (44.7% to 55.3%), with smaller proportions describing them as “completely capable” (5.3% to 7.9%). However, notable gaps in readiness were highlighted, with responses indicating “somewhat incapable” (18.4% to 26.3%) or “mostly incapable” (10.5% to 26.3%) in significant proportions. Scenarios involving problematic online buying/shopping and use of social media showed slightly higher incapability ratings compared to other cases, underscoring variability in perceived system preparedness. Figure 4 highlights the contrast between health systems capability and addiction workforce expertise.

**FIGURE 4.**
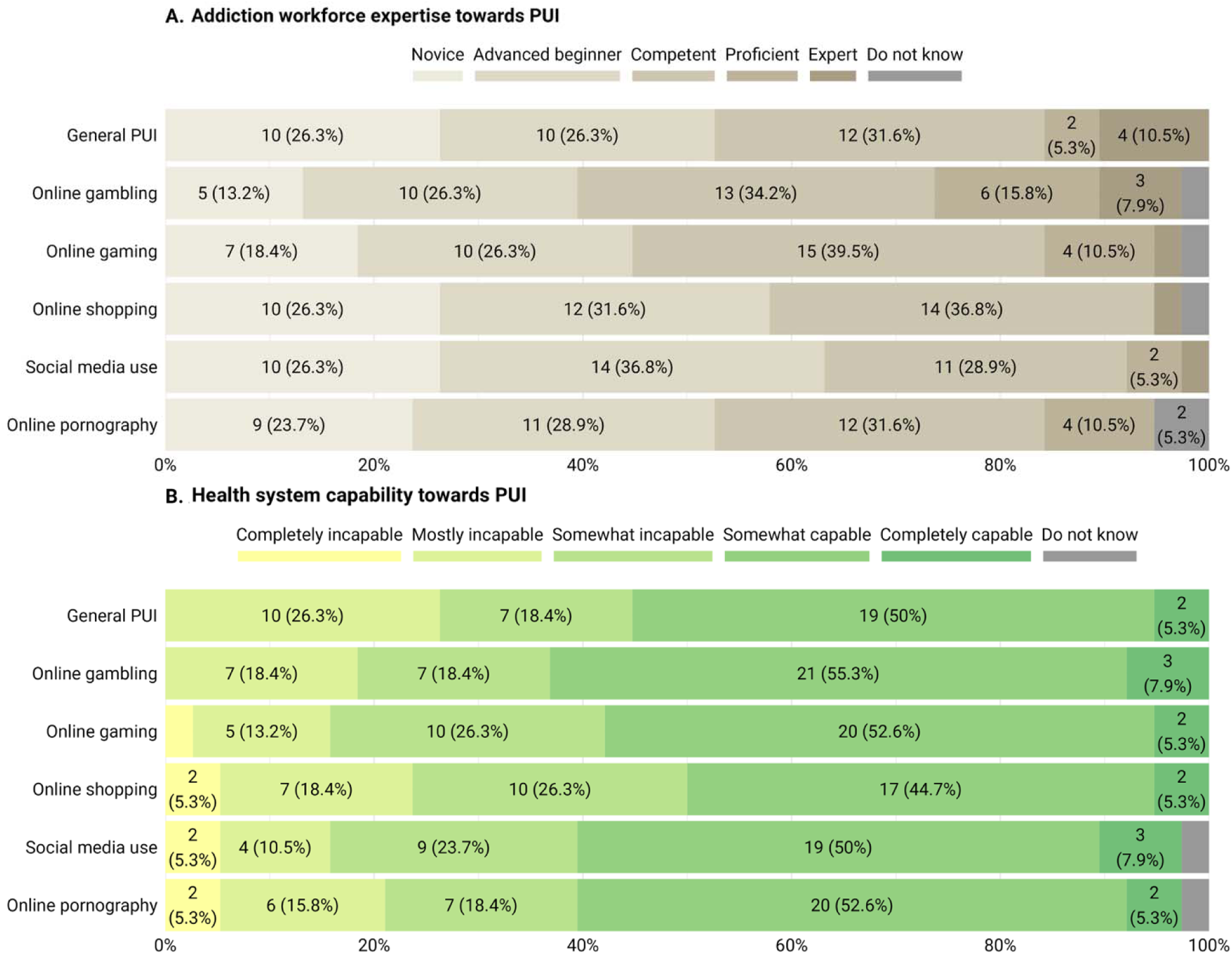
Health system capability in contrast to available addiction workforce expertise. figure (A) illustrates societies’ perspectives on the expertise level of their country’s addiction workforce in addressing PUI, ranging from novice to expert. Figure (B) presents their views on the capability of their country’s health system to manage PUI, measured on a scale from ‘completely incapable’ to ‘completely capable’.

Responses highlighted a mixed availability of distinct treatment and support programs for PUI within various countries’ health systems. While 2.6% (n = 1) of respondents indicated the presence of nationwide specific PUI treatment programs, an equal proportion of 44.7% (n = 17) reported non-nationwide programs or the absence of any programs. Expert awareness programs showed slightly better coverage, with 10.5% (n = 4) being nationwide and 39.5% (n = 15) available without nationwide reach, though a significant proportion of 42.1% (n = 16) noted their absence. Similarly, 2.6% (n = 1) of general educational programs were developed nationwide, while half (50.0%, n = 19) were available regionally. PUI prevention and screening programs were less commonly reported available between countries, with only 7.9% (n = 3) and 5.3% (n = 2), respectively, developed for nationwide use, while substantial gaps were noted as 50.0% (n = 19) and 68.4% (n = 26) of respondents reported no such programs. Supplementary Figure 2 highlights the general and country-specific availability of the aforementioned programs related to PUI.

Public awareness of PUI remained limited, with only 36.8% (n = 14) describing the community as adequately aware, while 39.5% (n = 15) rated awareness as partial. In contrast, addiction experts demonstrated higher levels of awareness, with 23.7% (n = 9) reporting them as fully aware and 47.4% (n = 18) as adequately informed, highlighting a disparity between professional and public understanding. Supplementary Figure 3 demonstrates this contrast between community and experts’ awareness levels in general and country-specific scales.

### PUI prioritization and engagement among societies

The survey results indicate varying levels of engagement and prioritization regarding PUI within countries and addiction medicine societies. Interest groups focused on PUI were reported by 44.7% (n = 17), while 24.2% (n = 13) noted their absence. Expert training programs on PUI were uncommon, with only 31.6% (n = 12) confirming their existence, compared to 63.2% (n = 24) reporting no such programs. Certifications in PUI were even rarer, available in only 10.5% (n = 4) of cases, with 78.9% (n = 30) indicating no certifications.

The need for research development on PUI was recognized as very or extremely important by a combined 68.4% (n = 26), with only 13.2% (n = 5) rating it as somewhat important or less. Current availability of investment plans for education and development in PUI as a clinical field was reported by 31.6% (n = 12), though 39.5% (n = 15) had no such plans and 28.9% (n = 11) were unsure. These findings underscore a current gap in infrastructure and initiatives for PUI but suggest an increasing acknowledgment of its future importance.

Regarding the perceived importance of PUI, 31.6% (n = 12) of respondents rated it as very important and 15.8% (n = 6) as extremely important at present. However, 28.9% (n = 11) considered it important and 21.1% (n = 8) only somewhat important. However, when evaluating its future significance in the next 10 years, the proportion of respondents rating PUI as very important increased to 42.1% (n = 16), with 23.7% (n = 9) seeing it as extremely important, highlighting growing concern over time. Figure 5 provides an overview of the perceived importance of PUI and the extent of engagement by addiction medicine societies in developing programs and facilities.

**FIGURE 5.**
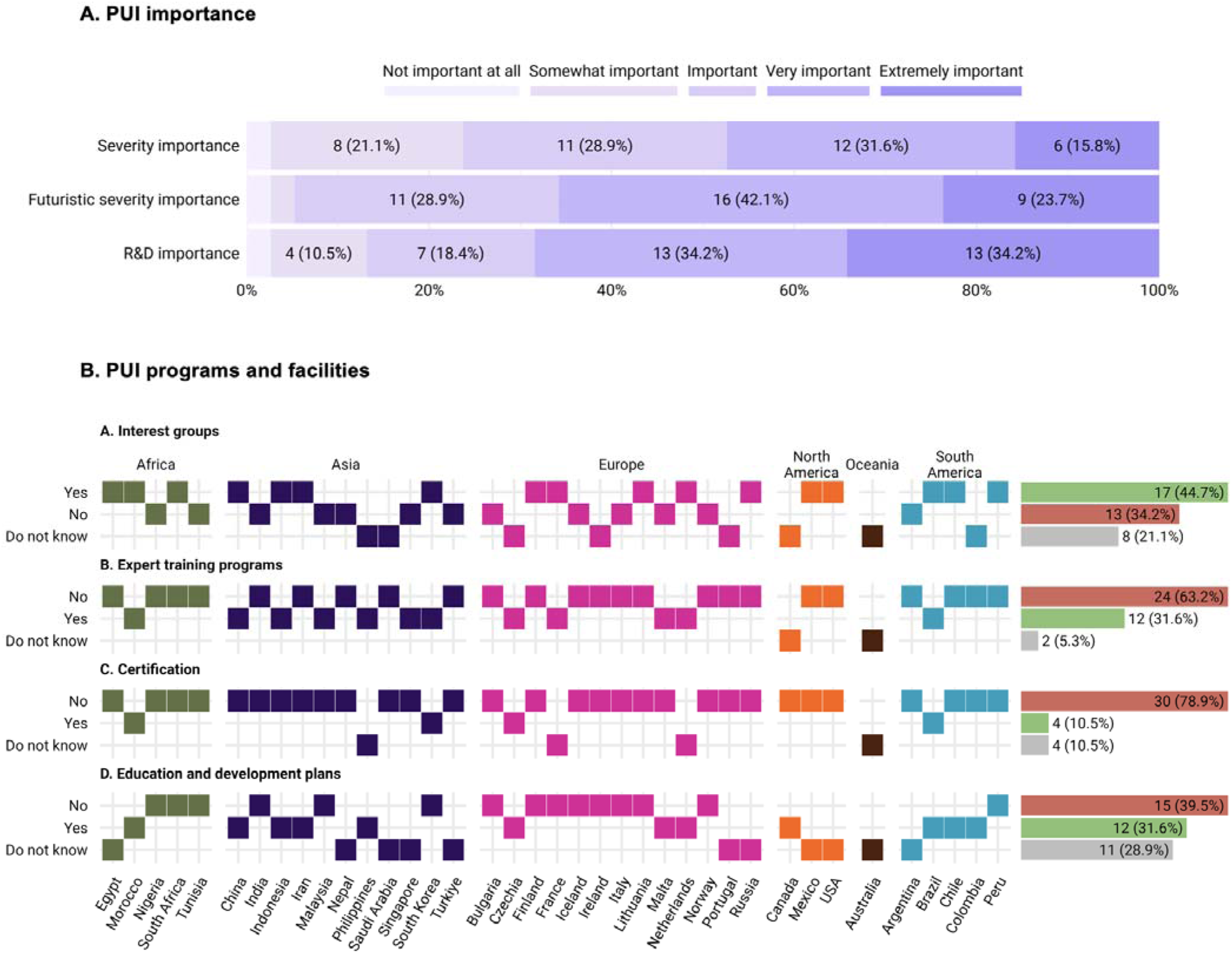
Importance of PUI and availability of supporting programs and facilities. Figure (A) illustrates societies’ views on the importance of PUI in three dimensions: its current severity, its projected severity over the next 10 years and its role in research development, rated on a scale from not important at all to extremely important. Figure (b) present perspectives on the availability of resources to support clinical responses to PUI, including interest groups, expert training programs, certification opportunities and investment in education and development.

## DISCUSSION

### Global relevance of and growing concerns regarding PUI

This international survey offers a global overiew on the health responses to PUI, showcasing a growing recognition of its significance alongside considerable gaps in infrastructure and preparedness. Among specific subtypes, problematic online gambling and gaming were most frequently reported, followed by social media use, pornography and online shopping. While psychotherapeutic interventions, particularly cognitive behavioral therapy, were widely available, the overall health response to PUI remains fragmented. Notably, the majority of surveyed societies emphasized the rising importance of PUI over the next decade, yet substantial deficiencies were identified in professional certification, practitioner education and expert training programs. These findings underscore a critical disparity between growing concern and actual readiness, highlighting the urgent need for coordinated global strategies to address PUI across clinical, educational and policy domains.

### Recognition of PUI’s severity and future projections

The findings of this global survey align with the increasing recognition of PUI as a important public health concern (8). Notably, epidemiological data on the prevalence of specific PUI subtypes remain scarce. This survey contributes valuable insights by highlighting the clinical presence of various PUI subtypes, each with distinct patterns of frequency, impact and severity. These variations underscore the complexity of PUI and need for subtype-specific research and interventions to address PUI’s multifaceted nature effectively (15, 16). The survey revealed widespread acknowledgment among addiction experts regarding the severity of PUI, with nearly half considering it extremely or very important at present. Future projections highlighted an even greater emphasis, with two-thirds of respondents anticipating PUI to remain a critical issue over the next decade. This growing recognition underscores the urgency for timely interventions and robust policy development.

PUI severity has been associated with mental health challenges and various psychological distress (17). Various factors may influence PUI severity, including sex (18–20), cultural differences and spirituality. For instance, current literature indicates that males are more vulnerable to PUI (21), particularly specific forms like those involving gaming (22), while emerging research explores the impact of cultural settings and spirituality on its severity (23, 24). Notably, this global survey revealed variations in PUI patterns across regions, with problematic use of online pornography and social media reported as milder in Asian countries compared to the Americas and Europe.

A significant portion of respondents also emphasized the importance of advancing PUI-related research and treatment development, reflecting the clinical importance of understanding and addressing this multifaceted phenomenon. These findings suggest the need for a globally coordinated approach to mitigate the impact of PUI effectively.

### Challenges in health system responses towards PUI

A key finding from the survey is the limitation of health systems to adequately address PUI. While nearly half of the respondents reporting the existence of some sort of treatment program in their country, only a small proportion indicated their availability on a nationwide scale. Aligning with existing literature, the most widely available options for tackling PUI rely on psychotherapeutic approaches such as CBT, mindfulness-based interventions and family or group therapy (25–27). However, there remains a notable lack of robust, context-specific data from various countries regarding the adaptation and effectiveness of these treatments for PUI, highlighting the need for targeted research and development in this area.

Despite PUI’s prevalence, no pharmacological treatments have been specifically approved for PUI, though our study suggests frequent off-label use of medications across different countries. This may stem from the frequent co-occurrence of PUI with psychiatric disorders such as depressive, anxiety, attention-deficit and other disorders, which often necessitate pharmacological management (28–30). Additionally, ongoing research into medications targeting compulsive behaviors and internet-related cravings provides a promising but as yet unestablished avenue for addressing PUI (31).

Awareness campaigns and prevention programs, which have shown some evidence of efficacy in similar contexts (32, 33), were reported to exist in limited capacities and remain far from universal implementation. This situation suggests significant gaps in health systems’ readiness and capacity to address PUI effectively (34). Strengthening these systems would benefit from global coordination to develop and assess standardized approaches and promote the exchange of successful strategies, paving the way for a more capable and unified response to this escalating public health concern.

### Engagement of addiction medicine/psychiatry societies

The role of addiction medicine societies in addressing PUI remains underdeveloped globally. While nearly half of respondents in this survey reported the presence of PUI-focused interest groups within their addiction medicine/psychiatry societies, only 10% indicated the availability of certification programs and less than one-third offered expert training programs. These findings underscore that while addiction experts are increasingly recognizing PUI as a critical issue, significant gaps persist in capacity building and resource allocation. A notable international effort under the European Cooperation in Science and Technology (COST) Action (CA 16207) established a global network of PUI researchers (EU-PUI) in 2018 (3). By the initiative’s conclusion in mid-2022, it had advanced the field through pivotal research on diagnosis, screening tools, underlying mechanisms and treatment approaches (11, 12, 35–37). Nevertheless, there is an urgent need for similar long-term initiatives at both national and international levels to sustain progress, enhance training and build infrastructure to address PUI effectively. Addiction medicine/psychiatry societies can take a more proactive stance by fostering interdisciplinary collaboration, advocating for resource allocation and developing standardized training and certification pathways.

### Challenges in awareness and public engagement

Our survey suggested a potential gap between the awareness of PUI among addiction experts and that within the general public, suggesting a possible barrier to effective prevention and early intervention efforts. Public engagement, particularly through patient and public involvement, offers a promising approach to bridging this divide by involving citizens at all stages of research and decision-making, thereby enhancing the quality and relevance of health initiatives (38, 39). Adapting this model, the EU-PUI COST Action has successfully illustrated the value of engaging diverse groups, including young people, parents, teachers and people with lived experience, to address knowledge gaps and community concerns (3). Key initiatives, such as international consultations and the International Festival of Science and Arts on PUI, have demonstrated public interest and highlighted critical challenges, such as the impact of PUI on families and the widening gap in technological literacy (40). Building on these successes requires the development of sustained frameworks for public engagement. Establishing advisory groups, co-designed training programs and collaborative platforms can empower citizens and promote shared ownership of PUI-related research and interventions. By integrating the public as active partners, health systems can cultivate a more informed and supportive environment, which is crucial for addressing the multifaceted challenges of PUI effectively.

### Study strengths and limitations

This study represents a comprehensive effort to capture the global landscape of PUI through the perspectives of addiction professionals. Its strengths include its wide geographical scope and the use of a validated global expert network, the ISAM-GEN and national addiction medicine/psychiatry societies to engage respondents. However, several limitations should be considered when interpreting the findings. First, the method used relied on an English-only survey, which may have excluded non-English-speaking professionals, potentially leading to language-based biases. Additionally, the non-responsive cohort could have skewed results, as certain regions or professional groups may be underrepresented. Furthermore, some important aspects, such as case scenarios involving multimorbidities, including those with both PUI and substance use disorders, were not addressed. The study also did not fully account for cultural and political contexts, which may influence the perception and management of PUI across different regions. Lastly, the narrow definition of treatment providers may have overlooked other potential contributors to PUI care. Future research could address these limitations by incorporating longitudinal designs, region-specific analyses and broader definitions of both treatment providers and PUI-related conditions.

### Call to action and policy implications

Addressing the global challenge of PUI requires a comprehensive, multi-faceted approach. Policymakers must prioritize the development of standardized diagnostic tools and context-specific interventions to improve prevention, treatment and research efforts. Health systems should expand their capacity by integrating PUI interventions into broader mental health and addiction services, ensuring accessibility even in resource-constrained settings. Collaboration at international, national and local levels is critical. Initiatives such as sustained global networks, public engagement frameworks and interdisciplinary training programs can enhance the field’s capacity to respond effectively. Policymakers and health authorities must also advocate for resource allocation, promote public awareness campaigns and support the active involvement of citizens in shaping interventions. A globally coordinated strategy will be pivotal in mitigating PUI’s impact and fostering a healthier digital landscape for future generations.

### Conclusion

This global survey provides valuable insights into the current landscape of PUI and highlights significant gaps in health response and opportunities in addressing this emerging public health concern. The findings underscore the growing recognition of PUI’s severity, variability in health system responses and underutilized role of addiction medicine/psychiatry societies. Despite increasing awareness among experts, the general public’s limited understanding poses challenges for prevention and intervention. Moving forward, coordinated global efforts are essential to bridge these gaps by developing standardized definitions, fostering capacity building and integrating PUI strategies into broader mental health frameworks. Collaborative initiatives and investments in research, education and policy development will be critical in addressing the multifaceted nature of PUI and mitigating its impact on global mental health.

### Declaration of generative AI and AI-assisted technologies in the writing process

During the preparation of this work, the authors used ChatGPT solely to improve English language fluency and grammar. After using this tool/service, the authors reviewed and edited the content as needed and take full responsibility for the content of the publication.

## Supporting information

Supplementary figures and full survey draft

## Data Availability

All data produced in the present study are available upon reasonable request to the authors.

## ACKNOWLEDGEMENTS

None

## Collaborators

ISAM-GEN Societies’ Experts: Wole Akosile^1^, Rabia Bilici^2^, Sari Castrén^3,4,5^, Steven KimWeng Chow^6^, Massimo Clerici^7^, Rui Manuel Vieira Gomes Correia^8^, Alessandra Diehl^9^, Nkereuwem William Ebiti^10^, Ali Farhoudian^11^, Dario Gigena Parker^12^, Jeffrey Gonzalez Giraldo^13^, Hugo González-Cantú^14^, Marie Grall-Bronnec^15,16^, Wei Hao^17^, Darius Jokūbonis^18,19^, Alexander Ivanov Kantchelov^20^, Emira Khelifa^21^, Evgeny Krupitsky^22^, Seung-Yup Lee^23,24^, David Robert Martell^25^, Garrett Gregory McGovern^26^, Belinda Julivia Murtani^27^, Valgerður Rúnarsdóttir^28^, Bigya Shah^29^, Wilco Sliedrecht^30,31^, Alireza Valyan^32^, Anna Maria Vella^33^, Salvador Benjamin Dimaya Vista^34^, Melvyn Weibin Zhang^35^

1. National Centre for Youth Substance Use Research, Faculty of Health and Behavioural Sciences, University of Queensland, Herston, Australia

2. Faculty of Medicine, Department of Psychology, Istanbul Ticaret University, Istanbul, Türkiye

3. Finnish Institute for Health and Welfare, Department of Public Health and Welfare, Helsinki, Finland

4. Social Sciences Department of Psychology and Speech-Language Pathology Turku, University of Turku, Turku, Finland

5. Department of Medicine, University of Helsinki, Helsinki, Finland

6. Addiction Medicine Association of Malaysia, Kuala Lumpur, Malaysia

7. Italian Society of Addiction Psychiatry (SIPDip), Milan, Italy

8. Associação Portuguesa da Medicina da Adição (APMA), Lisbon, Portugal

9. Brazilian Association for the Study of Alcohol and Other Drugs, Porto Alegre, Rio Grande do Sul, Brazil

10. Nigerian Society of Addiction Medicine, Nigeria

11. Department of Psychiatry, School of Medicine, Tehran University of Medical Sciences, Tehran, Iran

12. Public Health School, Faculty of Medical Science, National University of Cordoba, Cordoba, Argentina

13. Asociación Colombiana de Psiquiatría (ACP), Universidad del Rosario Bogotá-Colombia (CERSAME), Bogota, Colombia

14. International Society of Addiction Medicine, Calgary, Canada

15. Nantes Université, CHU Nantes, Institut Fédératif des Addictions Comportementales, F-44000 Nantes, France.

16. Nantes Université, Univ Tours, CHU Nantes, INSERM, MethodS in Patients centered outcomes and HEalth ResEarch, SPHERE, F-44000 Nantes, France.

17. Mental Health Institute, Central South University, Changsha, China

18. Lithuanian University of Health Sciences, Kaunas, Lithuania

19. Republican Center for Addictive Disorders, Vilnius, Lithuania

20. The Kantchelov Clinic, Sofiya, Bulgaria

21. Société Tunisienne d’addictologie, Faculty of medicine of Tunis, University Tunis el manar, Tunis, Tunisia

22. Russian Psychiatric Association, Russia

23. Department of Psychiatry, College of Medicine, The Catholic University of Korea, Seoul, Republic of Korea

24. Korean Academy of Addiction Psychiatry (KAAP), Republic of Korea

25. Canadian Society of Addiction Medicine, Calgary, Alberta, Canada

26. Irish Chapter of International Society of Addiction Medicine (IRE-ISAM), Dublin, Ireland

27. Addiction Psychiatry Section, Indonesian Psychiatric Association, Indonesia

28. SAA addiction medicine treatment centers, Iceland

29. Department of Psychiatry, School of Medicine, Patan Academy of Health Sciences, Nepal

30. Netherlands Society of Addiction Medicine, Netherlands

31. De Hoop foundation, Netherlands

32. Iranian National Center for Addiction Studies, Tehran University of Medical Sciences, Tehran, Iran

33. Foundation for Social Welfare Services (FSWS), Sedqa, Santa Venera, Malta

34. Philippine Addiction Specialists Society (PASS), Philippines

35. National Addictions Management Service, Institute of Mental Health, Singapore

---

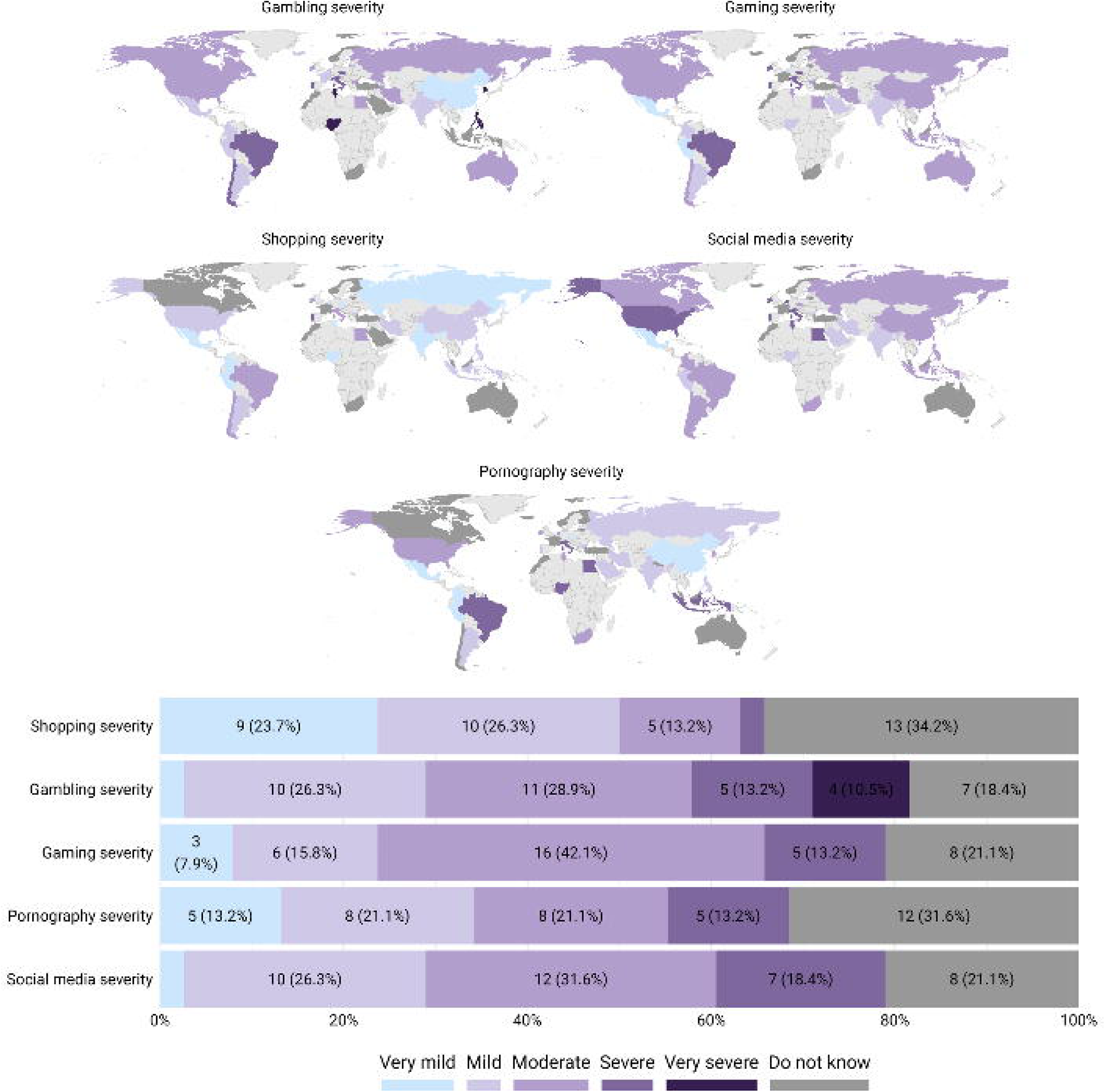

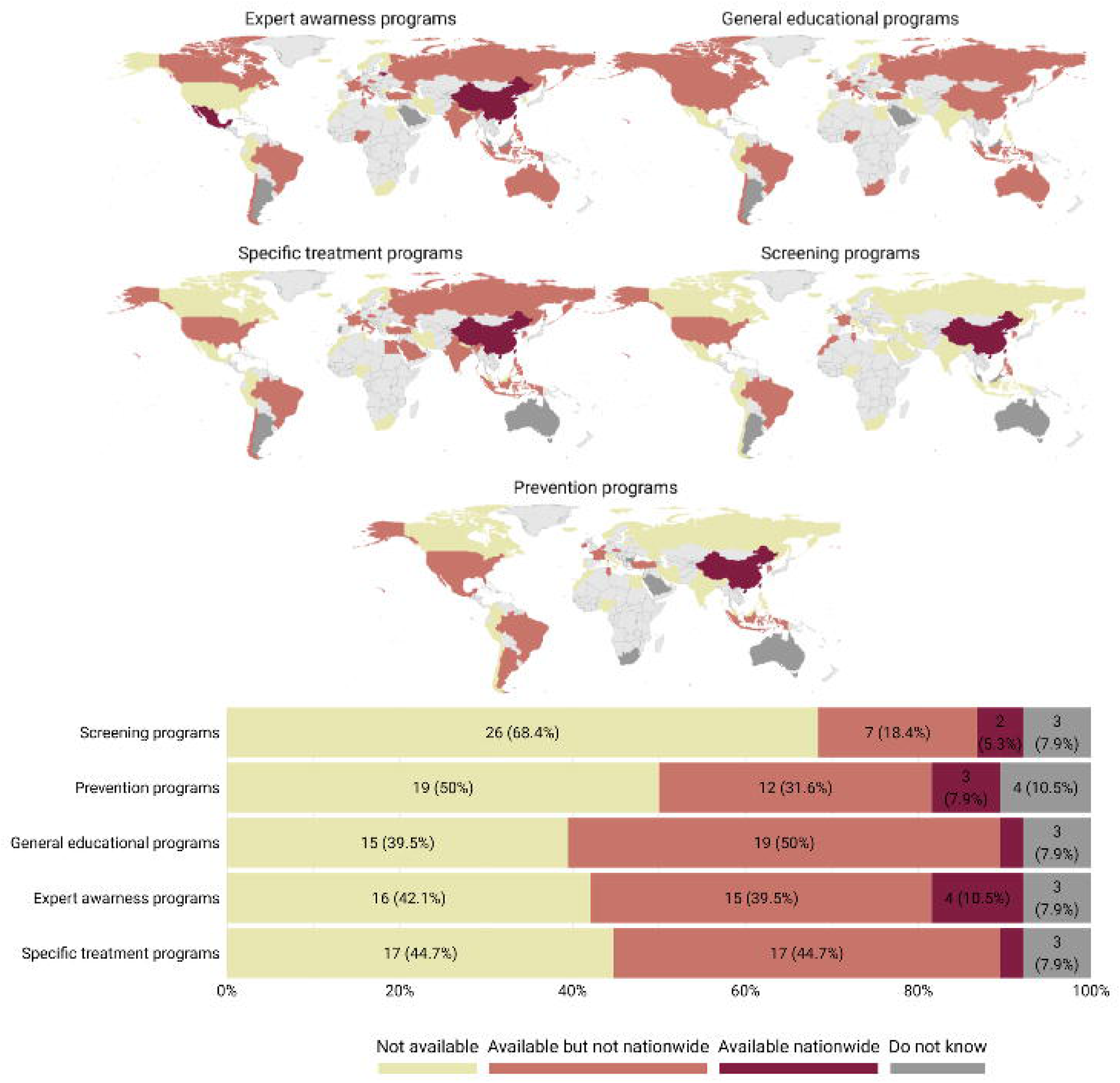

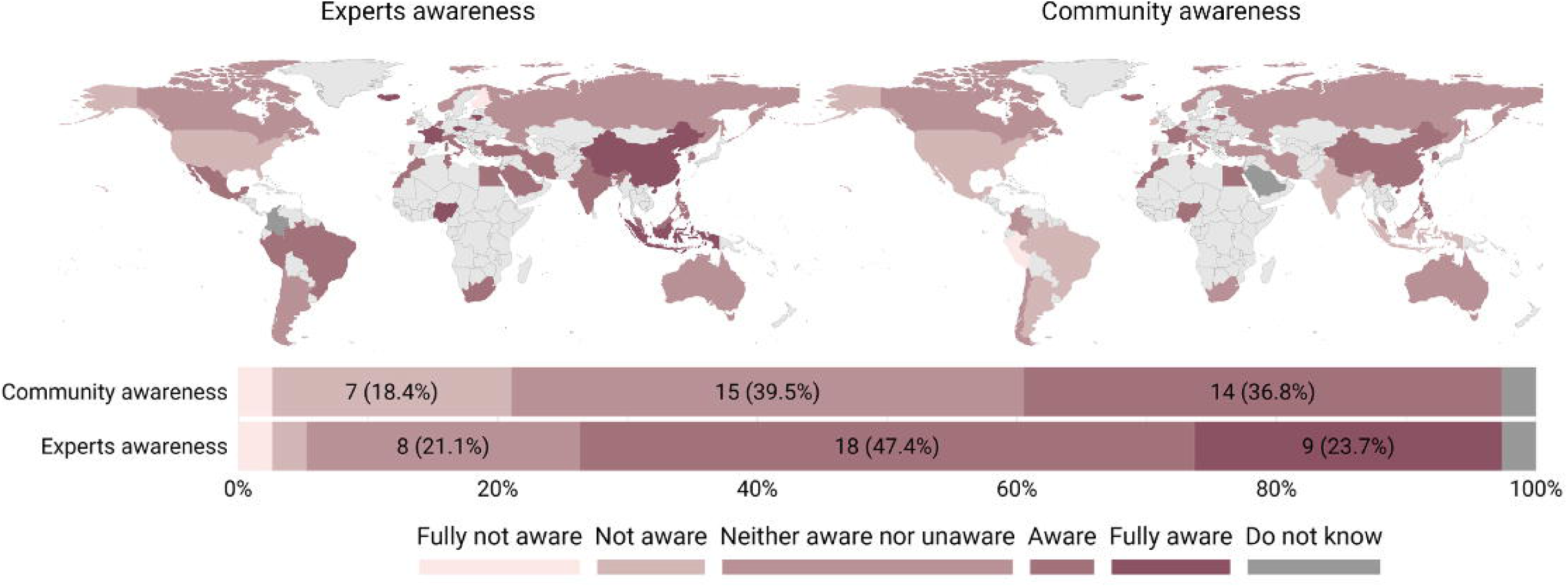

