## Supplementary figures and full survey draft for "Health Response to Problematic Usage of the Internet: A Global Survey on Trends, Available Treatments and Key Challenges"

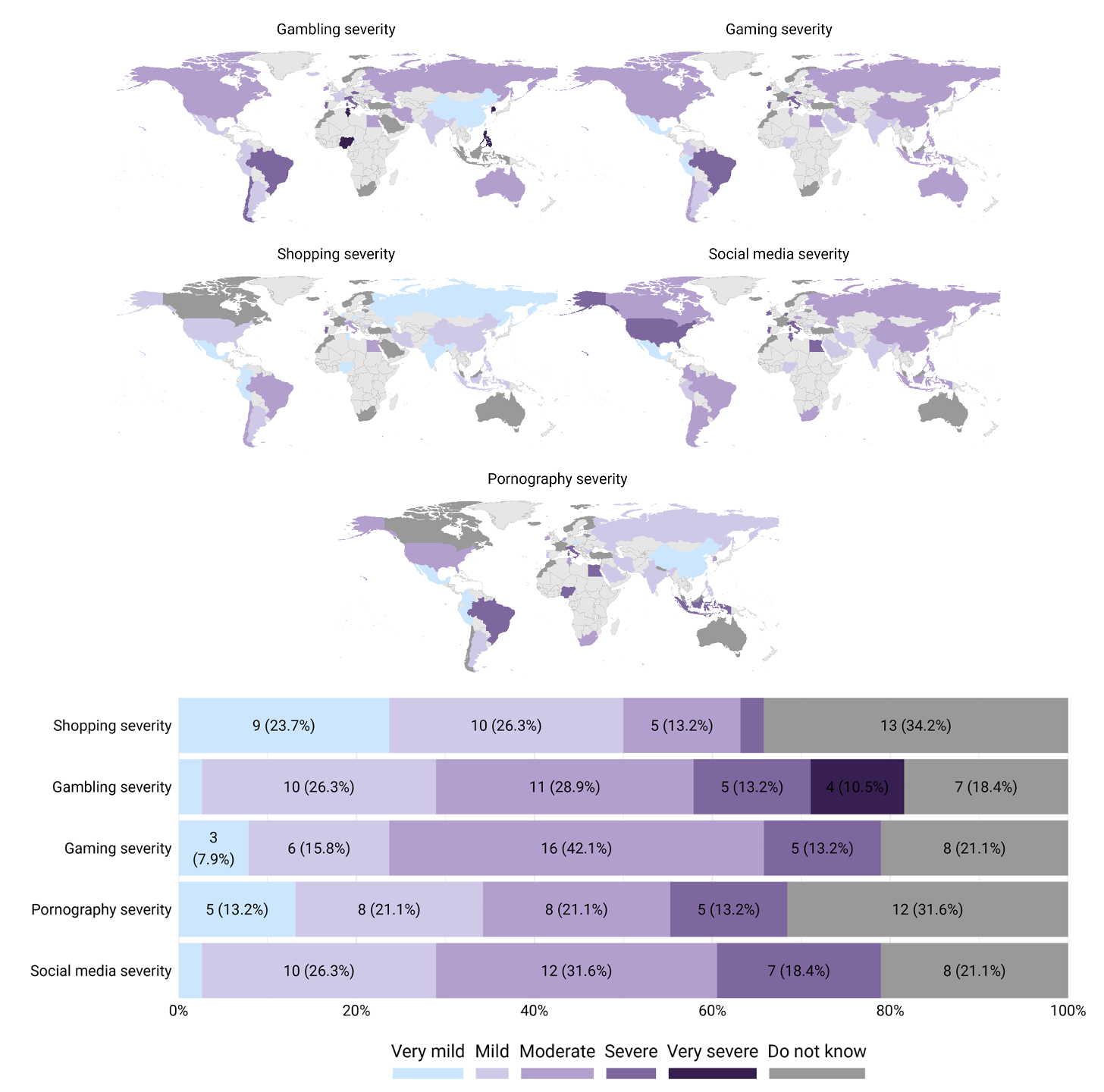

***Supplementary Figure 1. Severity of Distinct PUI Branches*** *– The five world heat maps provide a country-specific visualization of the severity of PUI branches, using the Likert scale to represent responses ranging from very mild to very severe. Additionally, the Likert scale diagram illustrates the severity of these distinct branches as reported by participating societies.*
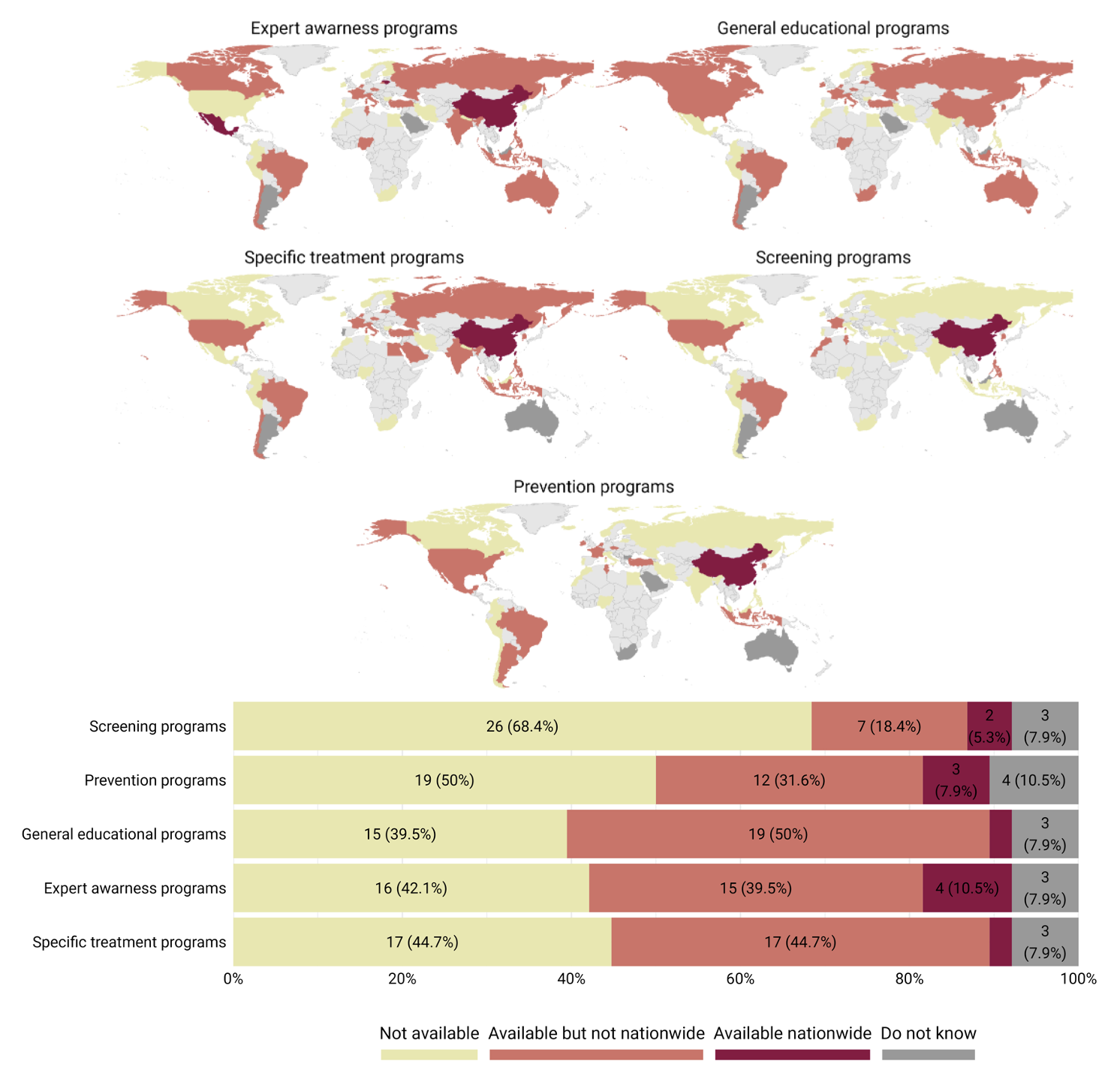

***Supplementary Figure 2. PUI-related Programs Availability Among the Countries*** *- The five world heat maps provide a country-specific visualization of the availability of programs for screening, prevention, general education, expert awareness, and specific PUI treatments, using the Likert scale to represent responses ranging from very mild to very severe. Additionally, the Likert scale diagram illustrates this generally as reported by participating societies.*

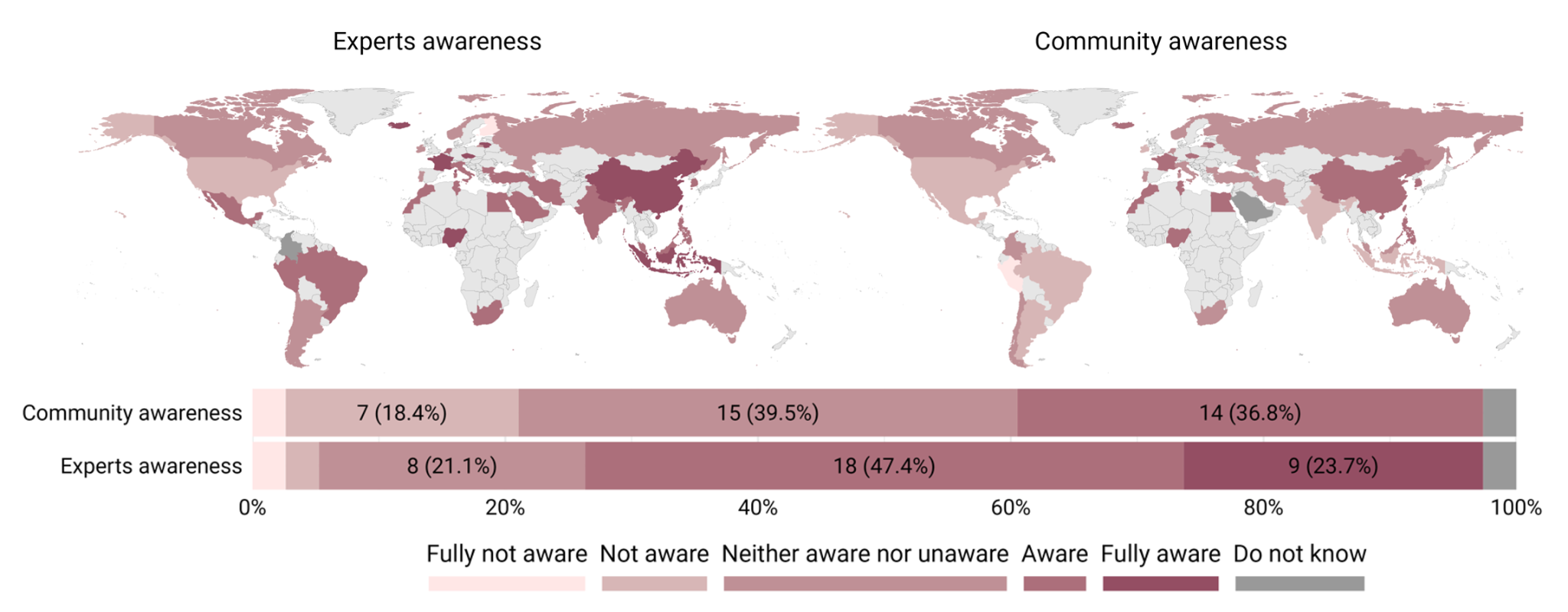

***Supplementary Figure 3. PUI Awareness Among the Community and Addiction Experts*** *- The two world heat maps provide a country-specific visualization of the awareness around PUI among addiction experts and the general community, using the Likert scale to represent responses ranging from fully not aware to fully aware. Additionally, the Likert scale diagram illustrates this awareness as reported by participating societies.*

**Full survey draft and raw responses:**

### Section 1. Demographic Information

1. First name
2. Middle name
3. Last name
4. Gender
   - Male ***(27, 71.05%)***
   - Female ***(11, 28.95%)***
   - Non-Binary/third gender ***(0, 0%)***
   - Prefer not to say ***(0, 0%)***
5. Age

- Under 25 ***(0, 0%)***
- 25-34 ***(1, 2.63%)***
- 35-44 ***(8, 21.05%)***
- 45-54 ***(12, 31.58%)***
- 55-64 ***(10, 26.32%)***
- 65 and over ***(7, 18.42%)***
- Prefer not to say ***(0, 0%)***

6. Academic degree *(You can select more than one option. Check all that apply.)*

- BSc or equivalents ***(0. 0%)***
- MSc or equivalents ***(2, 5.26%)***
- MD or equivalents ***(5, 13.16%)***
- MD, MSc or equivalents ***(7, 18.42%)***
- MD, PhD or equivalents ***(17, 44.74%)***
- PhD or equivalents ***(6, 15.79%)***
- Others, please specify …. ***(1, 2.63%)***

7. Name of your country, territory, region, or jurisdiction ***(Figure 1 of the main manuscript)***
8. Primary affiliation

- University / Teaching Hospital or Institution ***(20, 52.63%)***
- Independent Hospital ***(2, 5.26%)***
- Independent Research Institute ***(2, 5.26%)***
- Independent Clinical Institute/Center ***(4, 10.53%)***
- Government Health Program (such as Government ministry or department) ***(7, 18.42%)***
- Private Practice ***(3, 7.89%)***

9. Primary area of activity relevant to addiction

*Check all that apply.*

- Clinician ***(24, 63.16%)***
- Policy Maker ***(1, 2,63%)***
- Researcher/Scientist ***(8, 21.05%)***
- Administrator/Executive/Manager ***(4, 10.53%)***
- Others, please specify …. ***(1, 2.63%)***

10. Length of time (in years) in working with people with behavioral addictions directly or indirectly: ***Mean±SD = 20.71 ± 8.58***
11. Your position/designation within the society/organization
12. Email address
13. Affiliation

#### Responding on behalf of an addiction society/organization

Please consider a few points below before taking the survey on behalf of your addiction society/organization:

- The survey could be filled out by the society’s chair, president, or a delegated representative reflecting the society’s opinion and estimates. Meanwhile, the survey could also be shared within an internal group of experts selected by the society and the consensus will be reflected in the survey by the society’s representative.
- Since we are striving to collect this global data as precisely as possible, please provide your responses to the survey questions according to your society’s best estimate of the current state of your country.

14. Name of the society/organization you represent
15. Country/Territory/Region/Jurisdiction of the society/organization
16. Your position / designation in the society/organization
17. How many members does your society/organization have?

### Section 2 - PUI case scenario questions

Here we provide a series of case scenarios that describe how several PUI patients may present to addiction treatment services. Please answer the questions below of each scenario that ask how each patient might be managed in your country/jurisdiction.

Scenario 1 / 6. Alma is a 25-year-old who works as a software engineer in a multinational company. She spends most of her free time on the internet, browsing social media, playing online games, watching videos, and chatting with strangers. She often stays up late at night to log in to her favorite websites and neglects her sleep, hygiene, and health. She has lost interest in offline activities and hobbies that she used to enjoy before. She has also become distant from her family and friends, preferring to interact with her online contacts. She feels anxious and depressed when she is not online, and fantasizes about being on the internet when she is offline. Alma has tried to reduce her internet usage several times but failed to do so. She feels guilty and ashamed of her internet addiction but does not know how to stop it. She has also noticed that her work performance and productivity have significantly suffered because of her internet habits. She has received several warnings from her boss and colleagues about her poor quality of work and frequent mistakes. She fears that he might lose her job if she does not change her behavior. [General PUI Case]

1. In your region, this scenario is

- Non-existing ***(0, 0%)***
- Rare ***(0, 0%)***
- Uncommon ***(2, 5.26%)***
- Occasional ***(19, 50%)***
- Frequent ***(15, 39.47%)***
- Seen increasingly ***(0. 0%)***
- Do not know ***(2, 5.26%)***

2. The patient will most likely initially be seen by:

- General practitioner / Family doctor ***(15, 39.47%)***
- Emergency doctor ***(0, 0%)***
- Addiction medicine specialist physician ***(2, 5.26%)***
- Psychiatrist ***(6, 15.79%)***
- Psychologist/therapist/counselor ***(13, 34.21%)***
- Another health partitioner ***(0, 0%)***
- None of the above ***(2, 5.26%)***
- Do not know ***(0, 0%)***

3. The patient will most likely ultimately be/treated by:

- General practitioner / Family doctor ***(0, 0%)***
- Emergency doctor ***(0, 0%)***
- Addiction medicine specialist physician ***(9, 23.68%)***
- Psychiatrist ***(14, 36.84%)***
- Psychologist/therapist/counselor ***(14, 36.84%)***
- Another health partitioner ***(0, 0%)***
- None of the above ***(0, 0%)***
- Do not know ***(0, 0%)***

4. She will be treated with (you can select more than one):

- Pharmacological ***(16, 42.1%)***
- Neuromodulation (different brain stimulations including rTMS) ***(1, 2.6%)***
- Psychoeducation or education only ***(19, 50%)***
- CBT-based psychotherapy ***(26, 68.4%)***
- Mindfulness-based interventions ***(10, 26.3%)***
- Peer group support therapy ***(8, 21.1%)***
- Alternative medicine interventions (including homeopathy and acupuncture) ***(1, 2.6%)***
- System-level interventions (Family interventions, school, community, etc.) ***(6, 15.8%)***
- Other interventions (please specify) ***(8, 21.1%)***
- Do not know ***(0, 0%)***

5. She will most probably receive treatment in the following setting as the first line:

- Online short-term programs (up to one month) ***(1, 2.6%)***
- Outpatient short-term programs (up to one month) ***(7, 18.42%)***
- Residential short-term programs (up to one month) ***(0, 0%)***
- Inpatient (hospital-based) short-term programs (up to one month) ***(0, 0%)***
- Online long-term programs (more than one month) ***(1, 2.6%)***
- Outpatient long-term programs (more than one month) ***(25, 65.79%)***
- Residential long-term programs (more than one month) ***(1, 2.6%)***
- Inpatient (hospital-based) long-term programs (more than one month) ***(0, 0%)***
- Other setting (please specify) ***(2, 5.26%)***
- Do not know ***(1, 2.6%)***

6. How do you evaluate the expertise of the addiction workforce in confronting this scenario in your country?

- Novice ***(10, 26.32%)***
- Advanced beginner ***(10, 26.32%)***
- Competent ***(12, 31.58%)***
- Proficient ***(2, 5.26%)***
- Expert ***(4, 10.53%)***
- Do not know ***(0, 0%)***

7. How do you evaluate the capability of your country’s health system for managing such scenarios?

- Completely capable ***(2, 5.26%)***
- Somewhat capable **(19, 50%)**
- Somewhat incapable ***(7, 18.42%)***
- Mostly incapable ***(10, 26.32%)***
- Completely incapable ***(0, 0%)***
- Do not know ***(0, 0%)***

Scenario 2 / 6. Reza is a 23-year-old male who is obsessed with online gambling, especially sports betting. He spends most of his time on the internet, placing bets on various games and matches, hoping to win big. He often loses more money than he wins, but he cannot stop himself from chasing his losses despite his intention to do so. He has borrowed money from his friends and family and even resorted to illegal activities to fund his gambling habit while being warned by their sides as well. Reza feels sleep-deprived some days of the week and cannot make himself stop thinking about his gambling habits even when he is not online. He has lied to his loved ones about his gambling problem and has broken their trust and relationships. He feels hopeless and helpless about his situation but does not know how to quit gambling. He has also neglected his education and career and has dropped out of college. He has no goals or plans for his future and has isolated himself from society. [Online gambling case]

1. In your region, this scenario is

- Non-existing ***(1, 2.63%)***
- Rare ***(0, 0%)***
- Uncommon ***(1, 2.63%)***
- Occasional ***(12, 31.58%)***
- Frequent ***(24, 63.16%)***
- Seen increasingly ***(0. 0%)***
- Do not know ***(0, 0%)***

2. The patient will most likely initially be seen by:

- General practitioner / Family doctor ***(7, 18.42%)***
- Emergency doctor ***(0, 0%)***
- Addiction medicine specialist physician ***(3, 7.89%)***
- Psychiatrist ***(8, 21.05%)***
- Psychologist/therapist/counselor ***(18, 47.37%)***
- Another health partitioner ***(1, 2.63%)***
- None of the above ***(1, 2.63%)***
- Do not know ***(0, 0%)***

3. The patient will most likely ultimately be/treated by:

- General practitioner / Family doctor ***(0, 0%)***
- Emergency doctor ***(0, 0%)***
- Addiction medicine specialist physician ***(11, 28.95%)***
- Psychiatrist ***(13, 34.21%)***
- Psychologist/therapist/counselor ***(14, 36.84%)***
- Another health partitioner ***(0, 0%)***
- None of the above ***(0, 0%)***
- Do not know ***(0, 0%)***

4. He will be treated with (you can select more than one):

- Pharmacological ***(20, 52.6%)***
- Neuromodulation (different brain stimulations including rTMS) ***(4, 10.5%)***
- Psychoeducation or education only ***(16, 42.1%)***
- CBT-based psychotherapy ***(29, 76.3%)***
- Mindfulness-based interventions ***(11, 28.9%)***
- Peer group support therapy ***(15, 32.5%)***
- Alternative medicine interventions (including homeopathy and acupuncture) ***(1, 2.6%)***
- System-level interventions (Family interventions, school, community, etc.) ***(10, 26.3%)***
- Other interventions (please specify) ***(6, 15.8%)***
- Do not know ***(0, 0%)***

5. He will most probably receive treatment in the following setting as the first line:

- Online short-term programs (up to one month) ***(0, 0%)***
- Outpatient short-term programs (up to one month) ***(6, 15.79%)***
- Residential short-term programs (up to one month) ***(1, 2.63%)***
- Inpatient (hospital-based) short-term programs (up to one month) ***(2, 5.26%)***
- Online long-term programs (more than one month) ***(2, 5.26%)***
- Outpatient long-term programs (more than one month) ***(24, 63.16%)***
- Residential long-term programs (more than one month) ***(1, 2.6%)***
- Inpatient (hospital-based) long-term programs (more than one month) ***(0, 0%)***
- Other setting (please specify) ***(1, 2.63%)***
- Do not know ***(1, 2.63%)***

6. How do you evaluate the expertise of the addiction workforce in confronting this scenario in your country?

- Novice ***(5, 13.16%)***
- Advanced beginner ***(10, 26.32%)***
- Competent ***(13, 34.21%)***
- Proficient ***(6, 15.79%)***
- Expert ***(3, 7.89%)***
- Do not know ***(1, 2.63%)***

7. How do you evaluate the capability of your country’s health system for managing such scenarios?

- Completely capable ***(3, 7.89%)***
- Somewhat capable ***(21, 55.26%)***
- Somewhat incapable ***(7, 18.42%)***
- Mostly incapable ***(7, 18.42%)***
- Completely incapable ***(0. 0%)***
- Do not know ***(0. 0%)***

Scenario 3 / 6. Sara is a 19-year-old girl who lives with her aunt. She has been playing online role-playing games for the past four years, spending an average of eight hours per day on weekdays and up to 15 hours per day on weekends. She is obsessed with gaming and thinks about it constantly, even when she is not playing. She feels restless, irritable, and depressed when she is unable to access her games or when her aunt tries to limit her gaming time. She has developed a tolerance for gaming and needs to play more and more to achieve the same level of satisfaction. She has repeatedly attempted to quit or reduce her gaming time but always failed to do so. She has lost interest in other hobbies and activities that she used to enjoy, such as reading, painting, and going out with friends. She has continued to play games excessively despite knowing that it has caused her serious problems in her personal, academic, and occupational domains. She has lied to her aunt, her therapist, and her online friends about the amount of time she spends on gaming. She uses gaming as a way to escape from her negative emotions, such as loneliness, anxiety, and guilt. She has jeopardized or lost several opportunities and relationships because of her gaming behavior, such as dropping out of college, losing her part-time job, and breaking up with her boyfriend. [Online gaming case]

1. In your region, this scenario is

- Non-existing ***(0, 0%)***
- Rare ***(0, 0%)***
- Uncommon ***(3, 7.89%)***
- Occasional (***(15, 39.47%)***
- Frequent ***(18, 14.37%)***
- Seen increasingly ***(0, 0%)***
- Do not know ***(2, 5.26%)***

2. The patient will most likely initially be seen by:

- General practitioner / Family doctor ***(12, 31.58%)***
- Emergency doctor ***(0, 0%)***
- Addiction medicine specialist physician ***(2, 5.26%)***
- Psychiatrist ***(6, 15.79%)***
- Psychologist / therapist / counselor ***(15, 39.47%)***
- Another health partitioner ***(2, 5.26%)***
- None of the above ***(1, 2.63%)***
- Do not know ***(0, 0%)***

3. The patient will most likely ultimately be/treated by:

- General practitioner / Family doctor ***(1, 2.63%)***
- Emergency doctor ***(0, 0%)***
- Addiction medicine specialist physician ***(10, 26.32%)***
- Psychiatrist ***(12, 31.58%)***
- Psychologist/therapist/counselor ***(15, 39.47%)***
- Another health partitioner ***(0, 0%)***
- None of the above ***(0, 0%)***
- Do not know ***(0, 0%)***

4. She will be treated with (you can select more than one):

- Pharmacological ***(15, 39.5%)***
- Neuromodulation (different brain stimulations including rTMS) ***(1, 2.6%)***
- Psychoeducation or education only ***(18, 47.4%)***
- CBT-based psychotherapy ***(28, 73.7%)***
- Mindfulness-based interventions ***(13, 34.2%)***
- Peer group support therapy ***(12, 31.6%)***
- Alternative medicine interventions (including homeopathy and acupuncture) ***(2, 5.3%)***
- System-level interventions (Family interventions, school, community, etc.) ***(13, 34.2%)***
- Other interventions (please specify) ***(5, 13.2%)***
- Do not know ***(0, 0%)***

5. She will most probably receive treatment in the following setting as the first line:

- Online short-term programs (up to one month) ***(2, 5.26%)***
- Outpatient short-term programs (up to one month) ***(9, 23.68%)***
- Residential short-term programs (up to one month) ***(1, 2.63%)***
- Inpatient (hospital-based) short-term programs (up to one month) ***(1, 2.63%)***
- Online long-term programs (more than one month) ***(2, 5.26%)***
- Outpatient long-term programs (more than one month) ***(21, 55.26%)***
- Residential long-term programs (more than one month) ***(0, 0%)***
- Inpatient (hospital-based) long-term programs (more than one month) ***(0, 0%)***
- Other setting (please specify) ***(1, 2.63%)***
- Do not know ***(1, 2.63%)***

6. How do you evaluate the expertise of the addiction workforce in confronting this scenario in your country?

- Novice ***(7, 18.42%)***
- Advanced beginner ***(10, 26.32%)***
- Competent ***(15, 39.47%)***
- Proficient ***(4, 10.53%)***
- Expert ***(1, 2.63%)***
- Do not know ***(1, 2.63%)***

7. How do you evaluate the capability of your country’s health system for managing such scenarios?

- Completely capable ***(2, 5.26%)***
- Somewhat capable ***(20, 52.63%)***
- Somewhat incapable ***(10, 26.32%)***
- Mostly incapable ***(5, 13.16%)***
- Completely incapable ***(1, 2.63%)***
- Do not know ***(0, 0%)***

Scenario 4 / 6. Sam, aged 35, is a mechanical engineer working in a car manufacturing factory. . He has been significantly obsessed with buying the latest electronic gadgets, such as smartphones, laptops, tablets, cameras, and headphones online after the beginning of the COVID-19 pandemic. He often spends more than he can afford and uses multiple credit cards to pay for his online purchases. He has a large collection of gadgets that he rarely uses or even opens. He also neglects his work tasks, family, and social obligations because of his online shopping behavior. He feels anxious, restless, and irritable when he is not online shopping. He has received several warnings from his employer and his wife has threatened to divorce him because of his online shopping addiction. He recognizes that he has a problem, but he cannot stop or reduce his online shopping behavior. [Online shopping case]

1. In your region, this scenario is

- Non-existing ***(1, 2.63%)***
- Rare ***(6, 15.79%)***
- Uncommon ***(9, 23.68%)***
- Occasional ***(17, 44.74%)***
- Frequent ***(3, 7.89%)***
- Seen increasingly ***(0, 0%)***
- Do not know ***(2, 5.26%)***

2. The patient will most likely initially be seen by:

- General practitioner / Family doctor ***(10, 26.32%)***
- Emergency doctor ***(0, 0%)***
- Addiction medicine specialist physician ***(2, 5.26%)***
- Psychiatrist ***(7, 18.42%)***
- Psychologist / therapist / counselor ***(16, 42.11%)***
- Another health partitioner ***(1, 2.63%)***
- None of the above ***(1, 2.63%)***
- Do not know ***(1, 2.63%)***

3. The patient will most likely ultimately be/treated by:

- General practitioner / Family doctor ***(2, 5.26%)***
- Emergency doctor ***(0, 0%)***
- Addiction medicine specialist physician ***(7, 18.42%)***
- Psychiatrist ***(11, 28.95%)***
- Psychologist / therapist / counselor ***(16, 42.11%)***
- Another health partitioner ***(0, 0%)***
- None of the above ***(0, 0%)***
- Do not know ***(2, 5.26%)***

4. He will be treated with (you can select more than one):

- Pharmacological ***(13, 24.2%)***
- Neuromodulation (different brain stimulations including rTMS) ***(0, 0%)***
- Psychoeducation or education only ***(20, 52.6%)***
- CBT-based psychotherapy ***(27, 71.1%)***
- Mindfulness-based interventions ***(10, 26.3%)***
- Peer group support therapy ***(8, 21.1%)***
- Alternative medicine interventions (including homeopathy and acupuncture) ***(4, 10.5%)***
- System-level interventions (Family interventions, school, community, etc.) ***(6, 15.8%)***
- Other interventions (please specify) ***(5, 13.2%)***
- Do not know ***(2, 5.3%)***

5. He will most probably receive treatment in the following setting as the first line:

- Online short-term programs (up to one month) ***(0, 0%)***
- Outpatient short-term programs (up to one month) ***(13, 34.21%)***
- Residential short-term programs (up to one month) ***(0, 0%)***
- Inpatient (hospital-based) short-term programs (up to one month) ***(1, 2.63%)***
- Online long-term programs (more than one month) ***(3, 7.89%)***
- Outpatient long-term programs (more than one month) ***(17, 44.74%)***
- Residential long-term programs (more than one month) ***(0, 0%)***
- Inpatient (hospital-based) long-term programs (more than one month) ***(0, 0%)***
- Other setting (please specify) ***(2, 5.26%)***
- Do not know ***(2, 5.26%)***

6. How do you evaluate the expertise of the addiction workforce in confronting this scenario in your country?

- Novice ***(10, 26.32%)***
- Advanced beginner ***(12, 31.58%)***
- Competent ***(14, 36.84%)***
- Proficient ***(0, 0%)***
- Expert ***(1, 2.63%)***
- Do not know ***(1, 2.63%)***

7. How do you evaluate the capability of your country’s health system for managing such scenarios?

- Completely capable ***(2, 5.26%)***
- Somewhat capable ***(17, 44.74%)***
- Somewhat incapable ***(10, 26.32%)***
- Mostly incapable ***(7, 18.42%)***
- Completely incapable ***(2, 5.26%)***
- Do not know ***(0, 0%)***

Scenario 5 / 6. Chloe, a 26-year-old nurse, has always been fascinated by social media platforms. Over the past year, her engagement with these platforms has become increasingly problematic. She constantly checks her social media accounts during work hours, losing track of time and neglecting other tasks. Chloe now prioritizes social media over her once-beloved hobbies like hiking and painting, canceling plans with friends to stay home, and scrolling through her feeds. Despite negative consequences —such as poor work performance, sleep deprivation, and strained relationships— Chloe continues her social media use. She experiences significant distress and feels anxious when away from her phone or computer. This pattern has persisted for over a year, leaving her isolated and struggling to break free from the cycle. [Problematic social media use case]

1. In your region, this scenario is

- Non-existing ***(0, 0%)***
- Rare ***(1, 2.63%)***
- Uncommon ***(3, 7.89%)***
- Occasional ***(17, 44.74%)***
- Frequent ***(15, 39.47%)***
- Seen increasingly ***(0, 0%)***
- Do not know ***(2, 5.26%)***

2. The patient will most likely initially be seen by:

- General practitioner / Family doctor ***(11, 28.95%)***
- Emergency doctor ***(0, 0%)***
- Addiction medicine specialist physician ***(2, 5.26%)***
- Psychiatrist ***(4, 10.53%)***
- Psychologist / therapist / counselor ***(19, 50%)***
- Another health partitioner ***(1, 2.63%)***
- None of the above ***(1, 2.63%)***
- Do not know ***(0, 0%)***

3. The patient will most likely ultimately be/treated by:

- General practitioner / Family doctor ***(1, 2.63%)***
- Emergency doctor ***(0, 0%)***
- Addiction medicine specialist physician ***(6, 15.79%)***
- Psychiatrist ***(12, 31.58%)***
- Psychologist / therapist / counselor ***(18, 47.37%)***
- Another health practitioner ***(0, 0%)***
- None of the above ***(0, 0%)***
- Do not know ***(1, 2.63%)***

4. She will be treated with (you can select more than one):

- Pharmacological ***(8, 21.1%)***
- Neuromodulation (different brain stimulations including rTMS) ***(0, 0%)***
- Psychoeducation or education only ***(19, 50%)***
- CBT-based psychotherapy ***(30, 78.9%)***
- Mindfulness-based interventions ***(12, 31.6%)***
- Peer group support therapy ***(9, 23.7%)***
- Alternative medicine interventions (including homeopathy and acupuncture) ***(1, 2.6%)***
- System-level interventions (Family interventions, school, community, etc.) ***(4, 10.5%)***
- Other interventions (please specify) ***(5, 13.2%)***
- Do not know ***(2, 5.3%)***

5. She will most probably receive treatment in the following setting as the first line:

- Online short-term programs (up to one month)
- Outpatient short-term programs (up to one month) ***(15, 39.47%)***
- Residential short-term programs (up to one month) ***(0, 0%)***
- Inpatient (hospital-based) short-term programs (up to one month) ***(0, 0%)***
- Online long-term programs (more than one month) ***(2, 5.26%)***
- Outpatient long-term programs (more than one month) ***(17, 44.74%)***
- Residential long-term programs (more than one month) ***(0, 0%)***
- Inpatient (hospital-based) long-term programs (more than one month) ***(0, 0%)***
- Other setting (please specify) ***(2, 5.26%)***
- Do not know ***(2, 5.26%)***

6. How do you evaluate the expertise of the addiction workforce in confronting this scenario in your country?

- Novice ***(10, 26.32%)***
- Advanced beginner ***(14, 36.84%)***
- Competent ***(11, 28.95%)***
- Proficient ***(2, 5.26%)***
- Expert ***(1, 2.63%)***
- Do not know ***(0, 0%)***

7. How do you evaluate the capability of your country’s health system for managing such scenarios?

- Completely capable ***(3, 7.89%)***
- Somewhat capable ***(19, 50%)***
- Somewhat incapable ***(9, 23.68%)***
- Mostly incapable ***(4, 10.53%)***
- Completely incapable ***(2, 5.26%)***
- Do not know ***(1, 2.63%)***

Scenario 6 / 6. Griffin, a 30-year-old graphic designer, is experiencing a significant increase in the daily use of online pornography. Over the past year, his fixation on sexual content has spiraled out of control. His life orbits around explicit material, neglecting his design projects, hobbies, and even basic self-care. His partner feels distant, as Griffin prioritizes virtual encounters over their relationship. Despite installing filters, setting alarms, and being ashamed of the behavior, he remains ensnared, always finding a workaround to access online pornography content. His relationship is strained, marked by arguments and emotional distance. Legal repercussions loom after accidentally stumbling upon illegal material. Griffin's compulsive behavior has persisted for over a year, and shame and anxiety grip him daily. His work and family life quality significantly suffer, and he struggles to focus on anything unrelated to pornography. [Problematic online pornography use case]

1. In your region, this scenario is

- Non-existing ***(1, 2.63%)***
- Rare ***(1, 2.63%)***
- Uncommon ***(7, 18.42%)***
- Occasional ***(16, 42.11%)***
- Frequent ***(10, 26.32%)***
- Seen increasingly ***(1, 2.63%)***
- Do not know ***(2, 5.26%)***

2. The patient will most likely initially be seen by:

- General practitioner / Family doctor ***(7, 18.42%)***
- Emergency doctor ***(0, 0%)***
- Addiction medicine specialist physician ***(0, 0%)***
- Psychiatrist ***(10, 26.32%)***
- Psychologist / therapist / counselor ***(18, 47.37%)***
- Another health partitioner ***(0, 0%)***
- None of the above ***(2, 5.26%)***
- Do not know ***(1, 2.63%)***

3. The patient will most likely ultimately be/treated by:

- General practitioner / Family doctor ***(0, 0%)***
- Emergency doctor ***(0, 0%)***
- Addiction medicine specialist physician ***(4, 10.53%)***
- Psychiatrist ***(14, 36.84%)***
- Psychologist / therapist / counselor ***(18, 47.37%)***
- Another health practitioner ***(1, 2.63%)***
- None of the above ***(0, 0%)***
- Do not know ***(1, 2.63%)***

4. He will be treated with (you can select more than one):

- Pharmacological ***(11, 28.9%)***
- Neuromodulation (different brain stimulations including rTMS) ***(0, 0%)***
- Psychoeducation or education only ***(22, 57.9%)***
- CBT-based psychotherapy ***(31, 81.6%)***
- Mindfulness-based interventions ***(12, 31.6%)***
- Peer group support therapy ***(13, 34.2%)***
- Alternative medicine interventions (including homeopathy and acupuncture) ***(2, 5.3%)***
- System-level interventions (Family interventions, school, community, etc.) ***(5, 13.2%)***
- Other interventions (please specify) ***(3, 7.9%)***
- Do not know ***(1, 2.3%)***

5. He will most probably receive treatment in the following setting as the first line:

- Online short-term programs (up to one month) ***(0, 0%)***
- Outpatient short-term programs (up to one month) ***(9, 23.68%)***
- Residential short-term programs (up to one month) ***(1, 2.63%)***
- Inpatient (hospital-based) short-term programs (up to one month) ***(0, 0%)***
- Online long-term programs (more than one month) ***(2, 5.26%)***
- Outpatient long-term programs (more than one month) ***(23, 60.53%)***
- Residential long-term programs (more than one month) ***(0, 0%)***
- Inpatient (hospital-based) long-term programs (more than one month) ***(0, 0%)***
- Other setting (please specify) ***(2, 5.26%)***
- Do not know ***(1, 2.63%)***

6. How do you evaluate the expertise of the addiction workforce in confronting this scenario in your country?

- Novice ***(9, 23.68%)***
- Advanced beginner ***(11, 28.95%)***
- Competent ***(12, 31.58%)***
- Proficient ***(4, 10.53%)***
- Expert ***(0, 0%)***
- Do not know ***(2, 5.26%)***

7. How do you evaluate the capability of your country’s health system for managing such scenarios?

- Completely capable ***(2, 5.26%)***
- Somewhat capable ***(20, 52.63%)***
- Somewhat incapable ***(7, 18.42%)***
- Mostly incapable ***(6, 15.79%)***
- Completely incapable ***(2, 5.26%)***
- Do not know ***(1, 2.63%)***

### Section 3 - Evaluation of the importance of PUI on society/organization-level

Please respond to the questions in this section based on your response role as an individual expert from a country-level OR as a representative of a society/organization from the related society/organization-level.

1. Are there any interest groups available on PUI in your country OR society/organization?
   a. Yes ***(17, 44.74%)***
   b. No ***(13, 24.21%)***
   c. Do not know ***(8, 21.05%)***
2. Are there any specific expert training programs on PUI in your country OR society/organization?
   a. Yes ***(12, 31.58%)***
   b. No ***(24, 63.16%)***
   c. Do not know ***(2, 5.26%)***
3. Are there any specific certifications provided on PUI in your country OR society/organization?
   a. Yes ***(4, 10.53%)***
   b. No ***(30, 78.95%)***
   c. Do not know ***(4, 10.53%)***
4. How do you evaluate the importance of PUI from the perspective of your country OR society/organization in terms of severity at the moment?
   a. Not important at all ***(1, 2.63%)***
   b. Somewhat important ***(8, 21.05%)***
   c. Important ***(11, 28.95%)***
   d. Very important ***(12, 31.58%)***
   e. Extremely important ***(6, 15.79%)***
   f. Do not know ***(0, 0%)***
5. How do you evaluate the importance of PUI from the perspective of your country OR society/organization in terms of severity in the future (next 10 years)?
   a. Not important at all ***(1, 2.63%)***
   b. Somewhat important ***(1, 2.63%)***
   c. Important ***(11, 28.95%)***
   d. Very important ***(16, 42.11%)***
   e. Extremely important ***(9, 23.68%)***
   f. Do not know ***(0, 0%)***
6. How do you evaluate the importance of research development on PUI in your country OR society/organization?
   a. Not important at all ***(1, 2.63%)***
   b. Somewhat important ***(4, 10.53%)***
   c. Important ***(7, 18.42%)***
   d. Very important ***(13, 34.21%)***
   e. Extremely important ***(13, 34.21%)***
   f. Do not know ***(0, 0%)***
7. Do you have any investment plans for education and development of the PUI as a clinical field between addiction practitioners within your country OR society/organization for the future?
   a. Yes (please tell us more about this below) ***(12, 31.58%)***
   b. No ***(15, 39.47%)***
   c. Do not know ***(11, 28.95%)***
8. How do you evaluate your country's addiction experts OR your society/organization’s members' awareness of PUI?
   a. Fully aware (possesses proficiency and knowledge on the issue) ***(9, 23.68%)***
   b. Aware (can adequately understand the issue) ***(18, 47.37%)***
   c. Neither fully aware nor unaware (can understand some aspects of the issue) ***(8, 21.05%)***
   d. Not aware (can understand the issue only with the help of the experts) ***(1, 2.63%)***
   e. Fully not aware (can hardly understand the issue even with guidance from the experts) ***(1, 2.63%)***
   f. Do not know ***(1, 2.63%)***
9. To what extent are your country's addiction experts OR society/organization members active in terms of providing services related to PUI?
   a. Very active, they provide services related to PUI on a regular basis ***(8, 21.05%)***
   b. Somewhat active, they provide services related to PUI occasionally ***(16, 42.11%)***
   c. Not very active, they provide services related to PUI rarely ***(12, 31.58%)***
   d. Not active at all, they do not provide services related to PUI ***(2, 5.26%)***
   e. Do not know ***(0, 0%)***

### Section 4 - Country-level health response on PUI

### 1. Are there any specific treatment programs available for tackling issues related to PUI within the health system of your country?

a. Yes, it is developed and being used nationwide ***(1, 2.63%)***
b. Yes, but there are no nationwide programs available ***(17, 44.74%)***
c. No ***(17, 44.74%)***
d. Do not know ***(3, 7.89%)***

2. Are there any specific expert awareness programs available for PUI within the health system of your country?
   a. Yes, it is developed and being used nationwide ***(4, 10.53%)***
   b. Yes, but there are no nationwide programs available ***(15, 39.47%)***
   c. No ***(16, 42.11%)***
   d. Do not know ***(3, 7.89%)***
3. Are there any specific general educational programs available on PUI within the health system of your country?
   a. Yes, it is developed and being used nationwide ***(1, 2.63%)***
   b. Yes, but there are no nationwide programs available ***(19, 50%)***
   c. No ***(15, 39.47%)***
   d. Do not know ***(3, 7.89%)***
4. Are there any specific prevention programs available on PUI within the health system of your country?
   a. Yes, it is developed and being used nationwide ***(3, 7.89%)***
   b. Yes, but there are no nationwide programs available ***(12, 31.58%)***
   c. No ***(19, 50%)***
   d. Do not know ***(4, 10.53%)***
5. Are there any specific screening programs for PUI within the health system of your country?
   a. Yes, it is developed and being used nationwide ***(2, 5.26%)***
   b. Yes, but there are no nationwide programs available ***(7, 18.42%)***
   c. No ***(26, 68.42%)***
   d. Do not know ***(3, 7.89%)***
6. How do you evaluate community (public) awareness around PUI within the health system of your country?
   a. Fully aware (possesses proficiency and knowledge on the issue) ***(0, 0%)***
   b. Aware (can adequately understand the issue) ***(14, 36.84%)***
   c. Neither aware nor not aware (can understand some aspects of the issue) ***(15, 39.47%)***
   d. Not aware (can understand the issue only with the help of the experts) ***(7, 18.42%)***
   e. Fully not aware (can hardly understand the issue even with the guidance from the expert) ***(1, 2.63%)***
   f. Do not know ***(1, 2.63%)***
7. In general, how do you evaluate the availability of different treatment facilities for PUI within the health system of your country?

|  | Not available | Limitedly available | Somewhat available | Frequently available | Vastly available | Do not know | Prefer not to respond |
| --- | --- | --- | --- | --- | --- | --- | --- |
| Online short-term programs (up to one month) | ***(13, 34.21%)*** | ***(15, 39.47%)*** | ***(6, 15.79%)*** | ***(2, 5.26%)*** | ***(0, 0%)*** | ***(2, 5.26%)*** | ***(0, 0%)*** |
| Outpatient short-term programs (up to one month) | ***(6, 15.79%)*** | ***(13, 34.21%)*** | ***(11, 28.95%)*** | ***(4, 10.53%)*** | ***(2, 5.26%)*** | ***(2, 5.26%)*** | ***(0, 0%)*** |
| Residential short-term programs (up to one month) | ***(13, 34.21%)*** | ***(14, 36.84%)*** | ***(8, 21.05%)*** | ***(1, 2.63%)*** | ***(0, 0%)*** | ***(2, 5.26%)*** | ***(0, 0%)*** |
| Inpatient (hospital-based) short-term programs (up to one month) | ***(14, 36.84%)*** | ***(17, 44.74%)*** | ***(4, 10.53%)*** | ***(1, 2.63%)*** | ***(1, 2.63%)*** | ***(1, 2.63%)*** | ***(0, 0%)*** |
| Online long-term programs (more than one month) | ***(15, 39.47%)*** | ***(15, 39.47%)*** | ***(6, 15.79%)*** | ***(0, 0%)*** | ***(0, 0%)*** | ***(2, 5.26%)*** | ***(0, 0%)*** |
| Outpatient long-term programs (more than one month) | ***(4, 10.53%)*** | ***(14, 36.84%)*** | ***(12, 31.58%)*** | ***(4, 10.53%)*** | ***(3, 7.89%)*** | ***(1, 2.63%)*** | ***(0, 0%)*** |
| Residential long-term programs (more than one month) | ***(13, 34.21%)*** | ***(14, 36.84%)*** | ***(7, 18.42%)*** | ***(3, 7.89%)*** | ***(0, 0%)*** | ***(1, 2.63%)*** | ***(0, 0%)*** |
| Inpatient (hospital-based) long-term programs (more than one month) | ***(19, 50%)*** | ***(12, 31.58%)*** | ***(5, 13.16%)*** | ***(0, 0%)*** | ***(1, 2.63%)*** | ***(1, 2.63%)*** | ***(0, 0%)*** |
| Other settings (please specify) | ***(22, 57.89%)*** | ***(3, 7.89%)*** | ***(4, 10.53%)*** | ***(0, 0%)*** | ***(2, 5.26%)*** | ***(5, 13.16%)*** | ***(2, 5.26%)*** |

### Section 5 - Severity of distinct PUI types

1. How do you evaluate the severity of problematic online shopping as a PUI in your country/territory/jurisdiction?
   a. Very severe - more than 50% of online shoppers have problematic online shopping behavior ***(0, 0%)***
   b. Severe - between 30% and 50% of online shoppers have problematic online shopping behavior ***(1, 2.63%)***
   c. Moderate - between 10% and 30% of online shoppers have problematic online shopping behavior ***(5, 13.16%)***
   d. Mild - between 5% and 10% of online shoppers have problematic online shopping behavior ***(10, 26.32%)***
   e. Very mild - less than 5% of online shoppers have problematic online shopping behavior (***9, 23.68%)***
   f. Do not know ***(13, 34.21%)***
2. How do you evaluate the severity of problematic online gambling as a PUI in your country/territory/jurisdiction?
   a. Very severe - more than 50% of online gamblers have problematic online gambling behavior ***(4, 10.53%)***
   b. Severe - between 30% and 50% of online gamblers have problematic online gambling behavior ***(5, 13.16%)***
   c. Moderate - between 10% and 30% of online gamblers have problematic online gambling behavior ***(11, 28.95%)***
   d. Mild - between 5% and 10% of online gamblers have problematic online gambling behavior ***(10, 26.32%)***
   e. Very mild - less than 5% of online gamblers have problematic online gambling behavior ***(1, 2.63%)***
   f. Do not know ***(7, 18.42%)***
3. How do you evaluate the severity of problematic online gaming as a PUI in your country/territory/jurisdiction?
   a. Very severe - more than 50% of online gamers have problematic online gaming behavior ***(0, 0%)***
   b. Severe - between 30% and 50% of online gamers have problematic online gaming behavior ***(5, 13.16%)***
   c. Moderate - between 10% and 30% of online gamers have problematic online gaming behavior ***(5, 13.16%)***
   d. Mild - between 5% and 10% of online gamers have problematic online gaming behavior ***(6, 15.79%)***
   e. Very mild - less than 5% of online gamers have problematic online gaming behavior ***(3, 7.89%)***
   f. Do not know ***(8, 21.05%)***
4. How do you evaluate the severity of problematic online pornography as a PUI in your country/territory/jurisdiction?
   a. Very severe - more than 50% of people who use online pornography have problematic online pornography behavior ***(0, 0%)***
   b. Severe - between 30% and 50% of people who use online pornography have problematic online pornography behavior ***(5, 13.16%)***
   c. Moderate - between 10% and 30% of people who use online pornography have problematic online pornography behavior ***(8, 21.05%)***
   d. Mild - between 5% and 10% of people who use online pornography have problematic online pornography behavior ***(8, 21.05%)***
   e. Very mild - less than 5% of people who use online pornography have problematic online pornography behavior ***(5, 13.16%)***
   f. Do not know ***(12, 31.58%)***
5. How do you evaluate the severity of problematic online social media use as a PUI in your country/territory/jurisdiction?
   a. Very severe - more than 50% of online social media users have problematic online social media use behavior ***(0, 0%)***
   b. Severe - between 30% and 50% of online social media users have problematic online social media use behavior ***(7, 18.42%)***
   c. Moderate - between 10% and 30% of online social media users have problematic online social media use behavior ***(12, 31.58%)***
   d. Mild - between 5% and 10% of online social media users have problematic online social media use behavior ***(10, 26.32%)***
   e. Very mild - less than 5% of online social media users have problematic online social media use behavior ***(1, 2.63%)***
   f. Do not know ***(8, 21.05%)***

### Section 6 - Collecting further information on PUI

1. Please provide any additional relevant information, publications, or insights related to problematic Internet use from the point of view of your society/organization/country or your own self.
